# Peripheral TARC (CCL17) Levels Are Associated with Widespread Microstructural Vulnerability in Cognitively Unimpaired Older African Americans

**DOI:** 10.64898/2026.05.12.26353011

**Authors:** Soodeh Moallemian, Samira Raminfard, Isha Mhatre-Winters, Miray Budak, Bernadette A. Fausto, Jason R. Richardson, Mark A. Gluck

**Author notes:** Corresponding Author: Jason R. Richardson. These authors contributed equally: Jason R. Richardson, Mark A. Gluck.

## Abstract

Neuroinflammation is increasingly recognized as an early contributor to Alzheimer’s disease and related dementias (AD/ADRD), yet accessible blood-based biomarkers of early brain microstructural changes remain limited, particularly in African Americans, who are at elevated risk and underrepresented in biomarker research. We examined associations between plasma inflammatory markers and diffusion MRI measures in 41 cognitively unimpaired older African Americans (mean age, 68.7 years). Plasma chemokines were quantified using a multiplex immunoassay, and voxel-wise general linear models and cluster-based analyses were performed to evaluate associations with mean diffusivity (MD), radial diffusivity (RD), mean kurtosis (MK), and radial kurtosis (RK), adjusting for age, sex, and education. Higher plasma TARC/CCL17 levels were associated with widespread increases in MD and RD across gray and white matter, whereas Eotaxin-3/CCL26 showed similar but more spatially restricted associations. Associations with kurtosis metrics were limited, and neither marker was associated with global cognition measured by the Montreal Cognitive Assessment. These findings indicate that diffusivity measures are more sensitive than kurtosis metrics for detecting inflammation-related microstructural alterations. Peripheral chemokines, particularly TARC/CCL17, may serve as accessible biomarkers of preclinical brain microstructural vulnerability relevant to AD/ADRD risk in older African Americans.

## 1 Background

Converging evidence from genetics, molecular neuropathology, and neuroimaging implicates immune dysregulation as an implicated contributor of neural vulnerability in Alzheimer’s disease (AD) and AD-related dementias (AD/ADRD) (Heneka et al., 2025; Jack et al., 2018). Genome-wide association studies have identified immune-related genes-including TREM2, CD33, and complement pathway components as key modulators of AD risk (Bellenguez et al., 2022), while postmortem and in-vivo studies demonstrate early microglial activation preceding overt neurodegeneration (Hansen et al., 2018). Importantly, inflammatory processes appear to emerge decades before clinical symptom onset, suggesting that neuroimmune signaling may contribute to early microstructural brain alterations rather than simply reflecting downstream pathology (Banks, 2005; Racine et al., 2016).

Peripheral inflammation may influence central nervous system structure through multiple mechanisms, including blood-brain barrier modulation, cytokine-mediated microglial activation, and oligodendrocyte dysfunction (Franklin and ffrench-Constant, 2017; Zheng et al., 2016). Chemokines represent a special class of cytokines that regulate immune cell trafficking and activation during inflammatory responses. Several chemokines have been implicated in neurodegenerative processes and AD pathology. Thymus and activation-regulated chemokine (TARC/CCL17) has been reported to increase in AD and other inflammatory conditions(Campbell et al., 1999; Imai et al., 1998).

Similarly, elevated cerebrospinal fluid levels of C–C motif chemokine ligand 26 (CCL26; also known as eotaxin-3) have been reported in prodromal AD(Westin et al., 2012). However, the microstructural substrates linking systemic immune activity to early brain vulnerability remain poorly characterized. Diffusion-weighted imaging (DWI) provides sensitive, non-invasive quantitative measures of cellular organization by characterizing water diffusion in relation to biological barriers. While mean diffusivity (MD) reflects overall water mobility, mean kurtosis (MK) captures deviations from Gaussian diffusion reflecting microstructural complexity and radial diffusivity (RD) and radial kurtosis (RK) are particularly sensitive to myelin integrity and microstructural complexity respectively (Plank et al., 2025; Wen et al., 2019; Zhang et al., 2023). Because these diffusion metrics are influenced by tissue geometry, axonal organization, and microstructural complexity, the direction of their alterations may vary depending on the underlying biological processes and stage of pathology. Notably, increases in radial diffusivity have been linked to myelin disruption and inflammatory processes in both preclinical AD and related conditions (Chu et al., 2022; Dean et al., 2017; Racine et al., 2019), suggesting that diffusion MRI may capture early signatures of immune-mediated brain vulnerability.

Understanding these processes is particularly important in populations that are disproportionately affected by AD/ADRD yet underrepresented in biomarker research. African Americans are almost twice at risk of developing AD/ADRD compared to non-Hispanic whites(Barnes, 2022). Characterizing how systemic inflammation relates to early brain microstructural alterations in this population is critical for addressing health disparities and identifying modifiable risk pathways.

In the present study, we examined associations between plasma inflammatory markers and diffusion MRI metrics, including MD, MK, RD, and RK, in cognitively unimpaired older African Americans. We utilized both voxel-based and ROI-based approaches to characterize systemic inflammatory profiles. We identified robust associations between diffusion-derived measures of brain microstructure and two chemokines in particular: TARC and Eotaxin-3. Higher levels of these chemokines were consistently associated with increased MD and RD across both gray and white matter regions, suggesting reduced microstructural integrity. In contrast, associations with MK and RK, which are sensitive to tissue complexity and microstructural heterogeneity, were more regionally variable, indicating that the effects of systemic inflammation on brain microstructure may depend on local tissue organization and underlying biological processes

## 2 Methods and Materials

### 2.1 Participants

41 participants (29 females) were drawn from the longitudinal Pathways to Healthy Aging and Brain Health Alliance study at Rutgers University-Newark. The study was approved by the Rutgers Institutional Review Board (eIRB) and conducted in accordance with the principles of the 1975 Helsinki Declaration. Inclusion and exclusion criteria are brought in Appendix. Forty-one participants met the inclusion criteria. Cognitively unimpaired status was defined using education-adjusted Montreal Cognitive Assessment (MoCA) cutoffs validated for non-Hispanic Black individuals (Milani et al., 2018). Demographic and biomarker characteristics are summarized in Table 1.

**Table 1.** Data Descriptives. 41 participants (29 females/12 males) were included in the analysis.

|  |  |  |  |  | Skewness |  | Kurtosis |  |
|---|---|---|---|---|---|---|---|---|
|  | **Mean** | **SD** | **Minimum** | **Maximum** | **Skewness** | **SE** | **Kurtosis** | **SE** |
| **Age** | 68.683 | 5.6500 | 60 | 89 | 1.253 | 0.369 | 2.951 | 0.724 |
| **MoCA** | 23.439 | 3.0826 | 19 | 29 | 0.296 | 0.369 | -1.003 | 0.724 |
| **Education** | 14.000 | 2.1448 | 10 | 18 | 0.272 | 0.369 | -0.532 | 0.724 |
| **IL-10** | 0.341 | 0.2769 | 0.1014 | 1.687 | 3.262 | 0.369 | 13.737 | 0.724 |
| **IL-1β** | 0.158 | 0.0988 | 0.0327 | 0.447 | 1.363 | 0.369 | 1.854 | 0.724 |
| **IL-5** | 1.058 | 0.9138 | 0.1443 | 4.571 | 2.412 | 0.369 | 6.192 | 0.724 |
| **IL-6** | 1.999 | 1.4825 | 0.2323 | 7.601 | 2.059 | 0.369 | 4.924 | 0.724 |
| **IL-7** | 4.069 | 2.0846 | 1.0677 | 10.708 | 1.231 | 0.369 | 1.842 | 0.724 |
| **MCP-1** | 107.454 | 34.6952 | 50.7518 | 183.989 | 0.522 | 0.369 | -0.762 | 0.724 |
| **MIP-1α** | 32.448 | 64.4461 | 9.5856 | 432.200 | 6.262 | 0.369 | 39.768 | 0.724 |
| **TNF-α** | 1.517 | 0.6082 | 0.5818 | 3.360 | 0.970 | 0.369 | 0.996 | 0.724 |
| **TARC** | 66.910 | 55.3089 | 17.6258 | 326.939 | 3.045 | 0.369 | 12.026 | 0.724 |
| **Eotaxin-3** | 28.036 | 33.9758 | 8.1982 | 167.695 | 3.268 | 0.369 | 10.036 | 0.724 |

### 2.2 Plasma Inflammatory Biomarkers

Fasting EDTA plasma samples were thawed on ice and centrifuged at 10,000 × g for 5 minutes at 4°C. All samples were assayed in duplicate and blinded to imaging data.

Inflammatory markers (Eotaxin-3, IL-1β, IL-5, IL-6, IL-7, IL-10, MCP-1, MIP-1α, TARC, TNF-α) were quantified using a customized multiplex U-PLEX (catalog# K15067M-2) assay (Meso Scale Discovery, Rockville, MD). Samples were run at neat dilution. The plates were read on the MESO QuickPlex SQ 120 instrument and analyzed using the Discovery Workbench software. Concentrations were derived from an 8-point calibration curve using a 4-parameter logistic model with 1/y² weighting. Quality control (QC) was assessed by adding an in-house pooled plasma sample to each plate to measure the percentage coefficient of variance across plates. Inter-plate assay precision for the internal QC sample was as follows: Eotaxin-3 – 8%, IL-10 – 6.9%, IL-1β – 10%, IL-5 – 11.5%, IL-6 – 9.3%, IL-7 – 6.3%, MCP-1 – 9.4%, MIP-1α – 9.7%, TARC – 5.4%, and TNFα – 9.9%, indicating acceptable assay reliability.

### 2.3 MRI Acquisition

MRI data were acquired on a 3T Siemens Prisma scanner (32-channel head coil) at the Rutgers University Brain Imaging Center. High-resolution T1-weighted images were obtained using an MP-RAGE sequence (1 mm isotropic resolution; TR = 2300 ms; TE = 2.98 ms; TI = 900 ms). Diffusion-weighted images were acquired with 2.0 mm isotropic resolution (TR = 3400 ms; TE = 71 ms), including 7 b0 volumes and 126 diffusion-weighted volumes across three shells (b = 500, 1000, 2000 s/mm²). Detailed MRI acquisition and curation information are brought in supplementary document.

### 2.4 MRI Data Processing

#### 2.4.1 Anatomical Preprocessing

T1-weighted (T1w) images were corrected for intensity non-uniformity using N4 bias field correction (Tustison et al., 2010) implemented in ANTs 2.4.3 and used anatomical references throughout the workflow. Images were reoriented to AC-PC alignment and nonlinearly registered to the MNI152NLin2009cAsym template using symmetric normalization (SyN)(Avants et al., 2008). Brain extraction was performed using SynthStrip (Hoopes et al., 2022), and automated tissue segmentation was conducted using SynthSeg (Billot et al., 2023a, 2023b) implemented in FreeSurfer (version 7.3.1).

#### 2.4.2 Diffusion Preprocessing

Diffusion data were preprocessed using QSIPrep (version 1.1.1)(Cieslak et al., 2021), a robust and reproducible pipeline, which is built on Nipype (version 1.9.1) and follows the NiPreps framework. The pipeline integrates tools from FSL, MRtrix3, and ANTs within a standardized framework. Details are explained in supplementary materials.

#### 2.4.3 Diffusion Model Fitting and Metric Derivation

Diffusion tensor and kurtosis models were estimated using DIPY(Garyfallidis et al., 2014). For each subject and session, a diffusion kurtosis imaging (DKI) model was fit to the preprocessed multi-shell DWI data within a brain mask using the corresponding gradients tables.

Maps of MD, RD, MK, and RK were derived from the fitted model. All derived maps were visually inspected to ensure data quality. For group-level analysis, diffusion metric maps were spatially normalized to MNI space using SPM12 in MATLAB 2024b.

### 2.5 Statistical Analyses

#### 2.5.1 Whole-Brain Voxel-Wise Analyses

Voxel-wise associations between plasma inflammatory markers and diffusion metrics were examined using general linear models in SPM12.

Separate regression models were estimated for each diffusion maps and each inflammatory biomarker. In each model, the diffusion metric served as the dependent variable, with the inflammatory marker as the predictor of interest. Age, sex, and years of education were included as covariates of no interest. Continuous variables were mean-centered prior to analysis. GM and WM explicit masks—created as explained in Appendix—were used to assess tissue-specific effects in all analyses.

Statistical inference was performed using a voxel-defining threshold of p < .001 (uncorrected), followed by cluster-level family-wise error (FWE) correction at p < .05. We tested for positive associations between MD and RD and inflammatory markers, while a negative association was tested between MK and RK and inflammatory markers.

#### 2.5.2 Cluster-Based Analyses

To further characterize the direction and magnitude of significant voxel-wise effects, a secondary region-of-interest (ROI) analysis was performed based on clusters identified in the whole-brain analyses. Significant clusters from each voxel-wise model were extracted from the SPM-generated cluster maps and converted into binary masks.

For each subject and diffusion map, mean diffusion values were extracted within each cluster mask. These cluster-level mean values were then used in follow-up regression analyses, with the corresponding inflammatory marker as the predictor of interest, while controlling for age, sex, and years of education. From these models, standardized effect sizes and confidence intervals were estimated for each cluster.

## 3 Results

### 3.1 Whole-Brain Voxel-Wise

Among all markers tested, TARC and Eotaxin-3 demonstrated the most robust and spatially consistent associations with microstructural measures, surviving correction for multiple comparisons. In contrast, the remaining inflammatory markers showed more limited or non-significant effects at the whole-brain level. For completeness and transparency, results for all tested markers are provided in the Supplementary Materials, while the main Results section focuses on findings related to TARC and Eotaxin-3.

Anatomical localization of gray matter (GM) clusters was determined using Harvard-Oxford Cortical atlas, Julich Histological atlas and AAL3 atlas, WM tracts were identified using the JHU ICBM-DTI-81 atlas and JHU white matter tractography atlas(Oishi et al., 2008). Detailed peak coordinates and cluster statistics for all significant findings are provided in the Supplementary Materials. Tables A-1-10, in the supplementary materials, present the summary statistics and cluster peak coordinate information for each voxel-wise analysis within GM and WM.

No clusters survived cluster-level FWE correction for the association between mean and radial kurtosis and Eotaxin-3 in either gray or white matter indicating a lack of significant relationships with microstructural complexity.

Exploratory analyses of the remaining inflammatory markers revealed a heterogeneous pattern of associations with diffusion MRI metrics. No significant associations were observed for TNF-α, IL- 1β, IL-10, or MCP-1. In contrast, higher IL-5 levels were associated with increased MD, RD, and MK within localized gray matter regions, including the insula and occipital cortex, as well as the left superior longitudinal fasciculus (Supplementary Tables A11–A13). IL-6 demonstrated positive associations with MD and RD within bilateral frontal cortical regions (Supplementary Table A14), whereas IL-7 showed more widespread associations with increased MD and RD involving the temporal pole, hippocampus, middle frontal cortex, occipital cortex, and cerebellum (Supplementary Table A15 and Figure A1). Among these markers, MIP-1α exhibited the most extensive effects, showing widespread positive associations with MD across multiple cortical and subcortical gray matter regions and major white matter tracts (Supplementary Figure A2, Table A16), including the superior longitudinal fasciculus, inferior fronto-occipital fasciculus, corticospinal tract, anterior thalamic radiation, and posterior limb of the internal capsule.

#### 3.1.1 Mean Diffusivity and TARC

Within gray matter, higher plasma TARC levels were associated with widespread increases in mean diffusivity, indicating reduced microstructural integrity. Posterior cortical regions showed the most prominent associations, including the lateral occipital cortex, angular gyrus, and superior parietal lobule. Additional clusters were observed in medial regions, including the anterior cingulate and paracingulate cortex, as well as posterior midline structures such as the precuneus and posterior cingulate cortex. Frontal involvement included the superior and middle frontal gyri, encompassing premotor and supplementary motor areas. Temporal lobe effects extended across the inferior and middle temporal gyri, temporal pole, and fusiform cortex, with additional involvement of limbic structures including the parahippocampal gyrus and amygdala. Subcortical and cerebellar regions, including the thalamus and cerebellar Crus I/II, were also implicated. Figure 1 (Panel A) illustrates the spatial distribution of these effects in GM.

**Figure 1.**
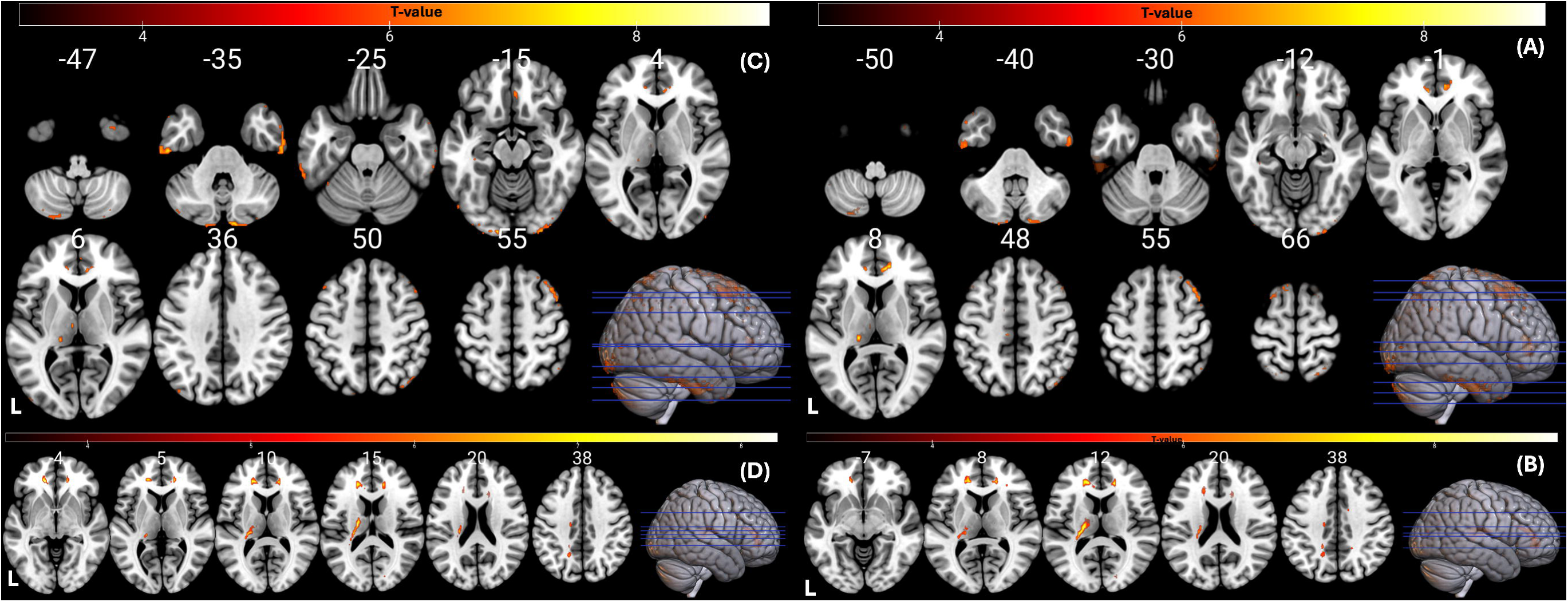
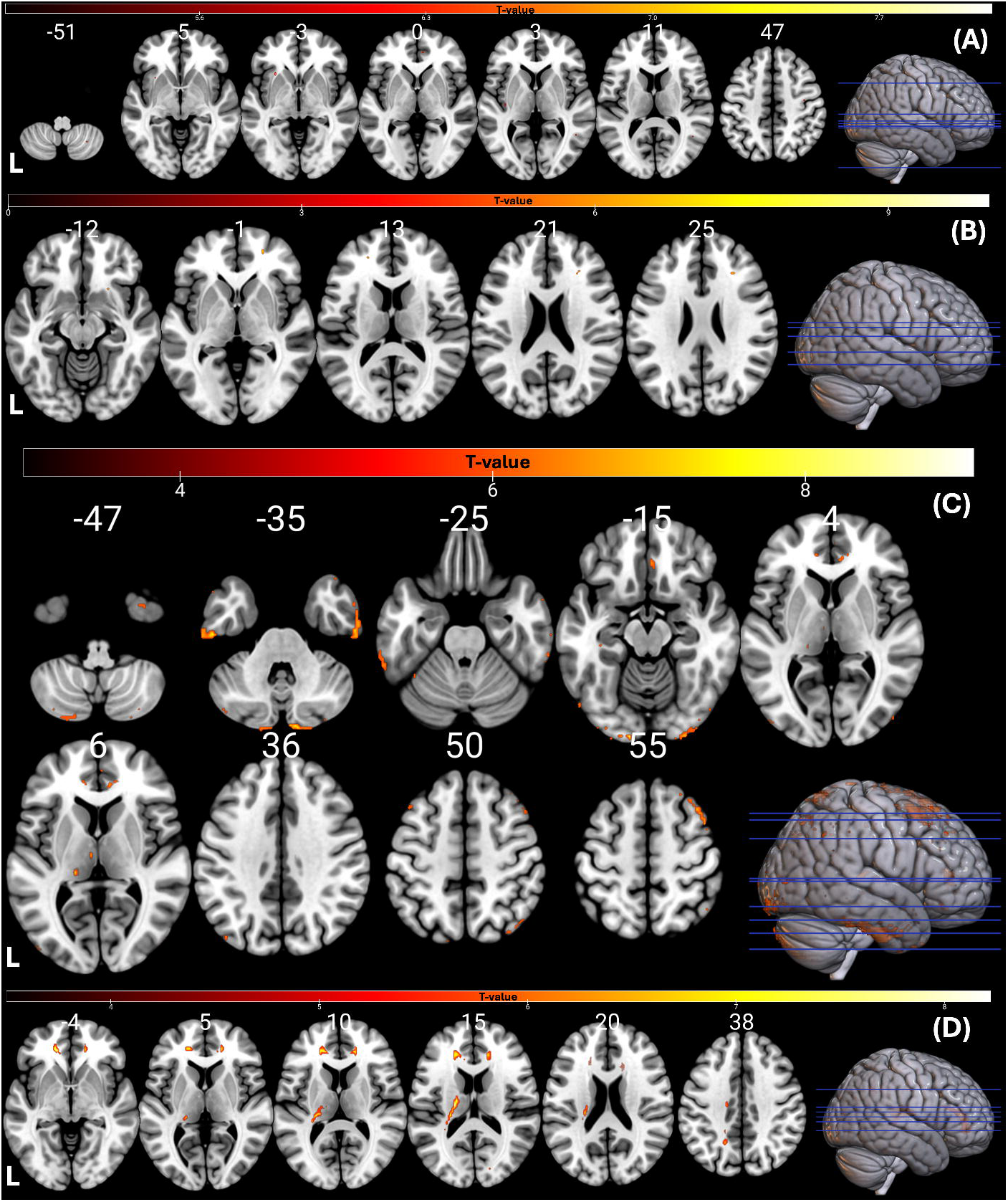
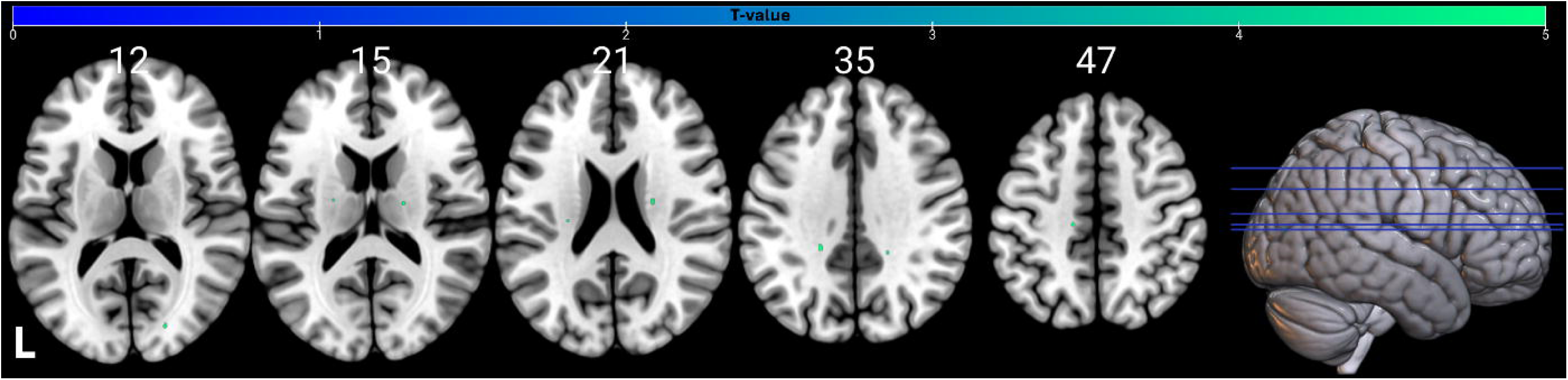
Statistical Parametric Maps. **MD signal increases with higher TARC levels in A) gray and B) white matter.** RD increases with higher TARC levels in C) gray and D) white matter. p_FWE_ <0 .05. T-value maps are overlaid on the MNI152 template. The color bar represents t-scores corresponding to the association with TARC. Abbreviations: MD, mean diffusivity; RD, radial diffusivity, FWE, family-wise error.

As presented in Figure 1 (panel B), in white matter, higher TARC levels were similarly associated with widespread increases in mean diffusivity, after FWER correction. The most prominent effects were observed in anterior white matter pathways, including the bilateral anterior corona radiata and forceps minor. Additional clusters were identified in major projection and association tracts, including the internal capsule, anterior thalamic radiation, and corticospinal tract. Posterior white matter regions were also involved, including the posterior corona radiata, posterior thalamic radiation, and occipital pathways such as the forceps major and inferior fronto-occipital fasciculus. Midline structures, including the corpus callosum and cingulum bundle, were consistently implicated.

#### 3.1.2 Radial Diffusivity and TARC

Within gray matter, higher TARC levels were associated with widespread increases in radial diffusivity in a spatial distribution closely mirroring that observed for mean diffusivity, spanning posterior cortical, medial, frontal, temporal, limbic, subcortical, and cerebellar regions (Figure 1, Panel C; peak coordinates in Supplementary Materials).

In white matter, higher TARC levels were similarly associated with widespread increases in radial diffusivity, paralleling the mean-diffusivity findings across the same anterior and posterior projection, commissural, and limbic pathways (Figure 1, Panel D).

#### 3.1.3 Mean Diffusivity and Eotaxin-3

In contrast to TARC, associations between Eotaxin-3 and mean diffusivity were more spatially restricted in both gray and white matter. Within gray matter, higher Eotaxin-3 levels were associated with increased mean diffusivity in a limited number of clusters surviving cluster-level FWER correction. Significant effects were observed in sensorimotor regions, including the precentral and postcentral gyri, as well as in frontal and limbic regions, including the frontal pole and temporal pole with extension into the parahippocampal gyrus and amygdala. Additional clusters were identified in posterior regions, including the inferior parietal lobule, as well as in cerebellar structures. Figure 2 (Panel A) illustrates the spatial distribution of these effects in GM.

**Figure 2.**
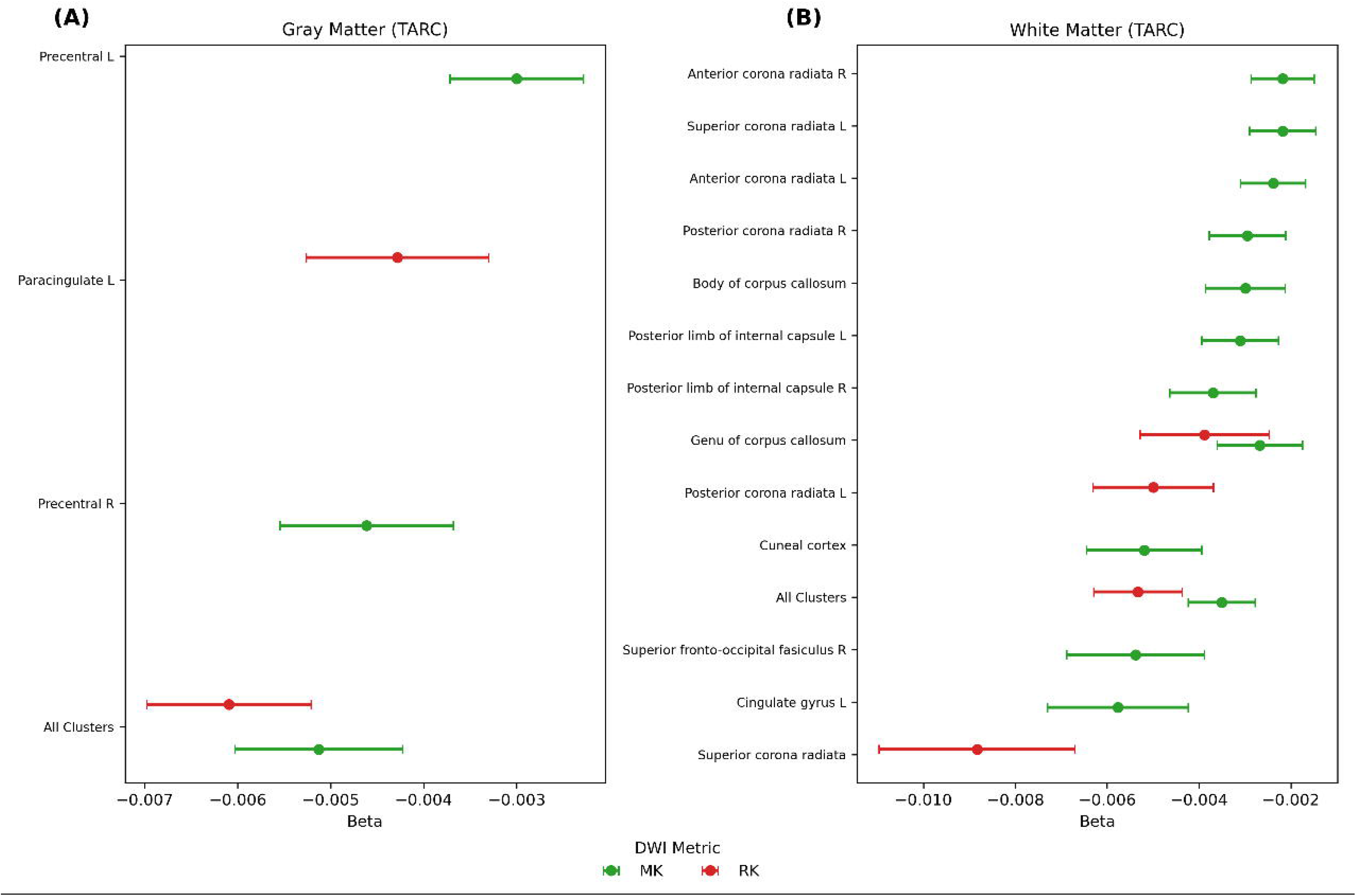
Statistical Parametric Maps. **MD and RD signal increase with higher Eotaxin-3 levels in gray and white matter.** Statistical parametric maps identify regions within (A) gray matter and (B) white matter where MD values increase with higher Eotaxin-3 levels (p_FWE_<0 .05). Panels (C) and (D) show the regions within GM and WM where RD increase with higher Eotaxin-3. T-value maps are overlaid on the MNI152 template. The color bar represents t-scores corresponding to the association with Eotaxin-3. Abbreviations: MD, mean diffusivity; RD, radial diffusivity; FWE, family-wise error.

Within white matter, Eotaxin-3-related increases in mean diffusivity were similarly localized. Significant clusters were primarily observed in anterior white matter pathways, including regions corresponding to the anterior thalamic radiation and inferior fronto-occipital fasciculus. Figure 2 (Panel B) shows the corresponding statistical parametric maps in WM.

#### 3.1.4 Radial Diffusivity and Eotaxin-3

Within gray matter, radial-diffusivity effects mirrored the mean-diffusivity distribution, involving sensorimotor, frontal, and limbic regions with smaller posterior parietal and cerebellar clusters (Figure 2, Panel C).

Within white matter, Eotaxin-3-related increases in radial diffusivity were minimal and restricted to a single small cluster in anterior regions, with limited anatomical specificity close to the right middle frontal gyrus with the cluster peak at (27, 30, 25). Figure 2 (Panel D) shows the corresponding statistical parametric map in WM.

#### 3.1.5 Mean Kurtosis and TARC

Associations between TARC and mean kurtosis were more spatially restricted than those observed for diffusivity-based metrics and showed distinct patterns across gray and white matter.

Within gray matter, higher TARC levels were associated with limited alterations in mean kurtosis, with only two small clusters localized to sensorimotor and posterior regions, including the precentral gyrus and adjacent medial parietal areas, as well as occipital regions encompassing the calcarine and precuneus.

After correcting for the FWER at voxel-level, only two voxels within each cluster, survived. In contrast, white matter exhibited more consistent and spatially distributed associations. Higher TARC levels were associated with significant alterations in mean kurtosis across major projection and commissural pathways, including the internal capsule, corona radiata, and corpus callosum. Additional involvement was observed in anterior white matter regions, including the anterior corona radiata and fronto-occipital pathways. Figure 3 shows the corresponding statistical parametric maps in WM.

**Figure 3.**
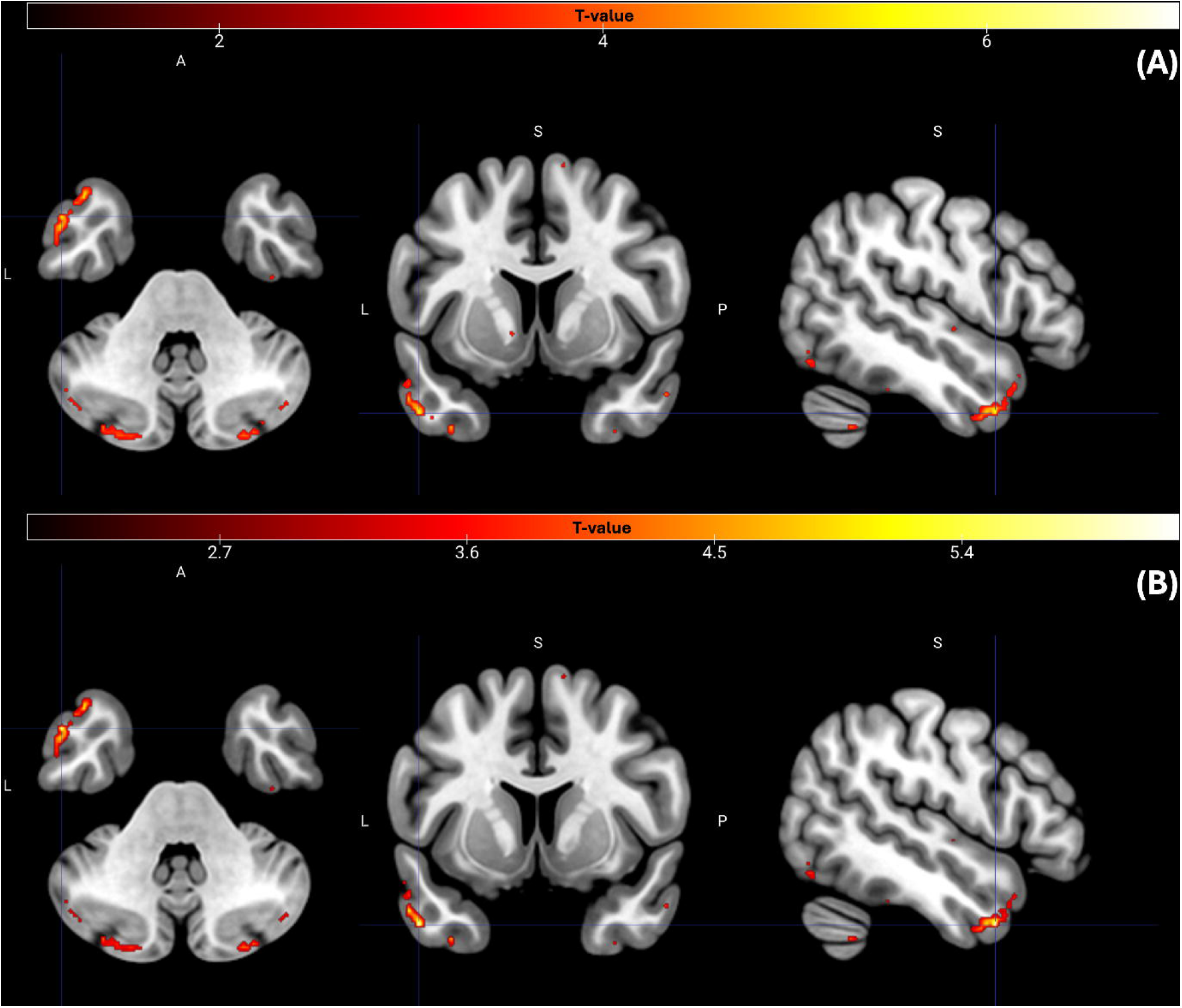
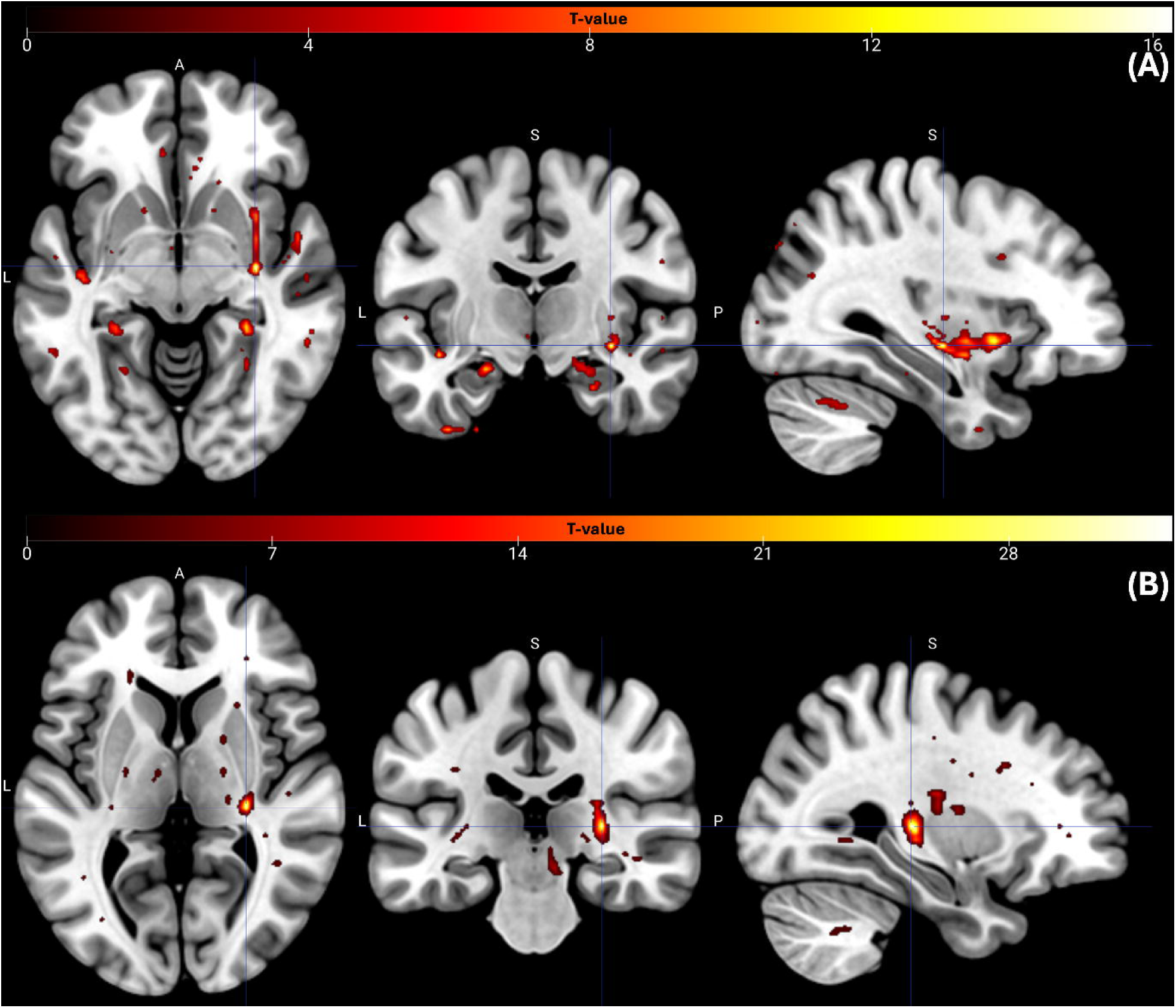
Statistical Parametric Maps. **White matter MK decreases with higher TARC levels.** Statistical parametric maps identify regions within white matter where MK values decrease with higher TARC levels (p_FWE_<0 .05). The t-value map is overlaid on the MNI152 template. The color bar represents t-scores corresponding to the association with TARC. Abbreviations: MK, mean kurtosis; FWE, family-wise error.

#### 3.1.6 Radial Kurtosis and TARC

Associations between TARC and radial kurtosis were limited and spatially restricted compared to those observed for diffusivity-based metrics.

Within gray matter, only a single small cluster survived cluster-level FWER correction at cluster-level, localized to the precentral region extending into medial parietal areas, however, this cluster did not survive FWER correction at voxel-level.

In white matter, a small number of clusters demonstrated significant associations, primarily within midline and projection pathways. These included the genu of the corpus callosum, as well as regions corresponding to the corona radiata and corticospinal tract. These effects did not survive the FWER correction at voxel-level.

### 3.2 Cluster-Based

Cluster-based ROI analyses were performed using masks derived from significant clusters (pFWE<0.05 at cluster-level) identified in the whole brain analyses. Tables A17-A28 present the cluster-based ROI analyses parameters.

#### 3.2.1 TARC

Cluster-based analyses confirmed robust associations between plasma TARC levels and diffusion MRI measures in both gray and white matter. In gray matter, higher TARC levels were consistently associated with increased MD and RD across multiple cortical and subcortical regions, with the strongest effects observed in temporal, parietal, and occipital areas. Similar positive associations were observed in major white matter pathways, including the corona radiata, internal capsule, thalamic radiations, and corpus callosum. In contrast, kurtosis metrics (MK and RK) demonstrated weaker and less consistent associations, with effect estimates generally centered near zero (Supplementary Figure A3). Figure 4, panels A and B, shows the standardized regression coefficients (β) and 95% confidence intervals for the association between plasma TARC levels and diffusion MRI metrics (MD and RD) extracted from significant clusters identified in voxel-wise analyses.

**Figure 4.**
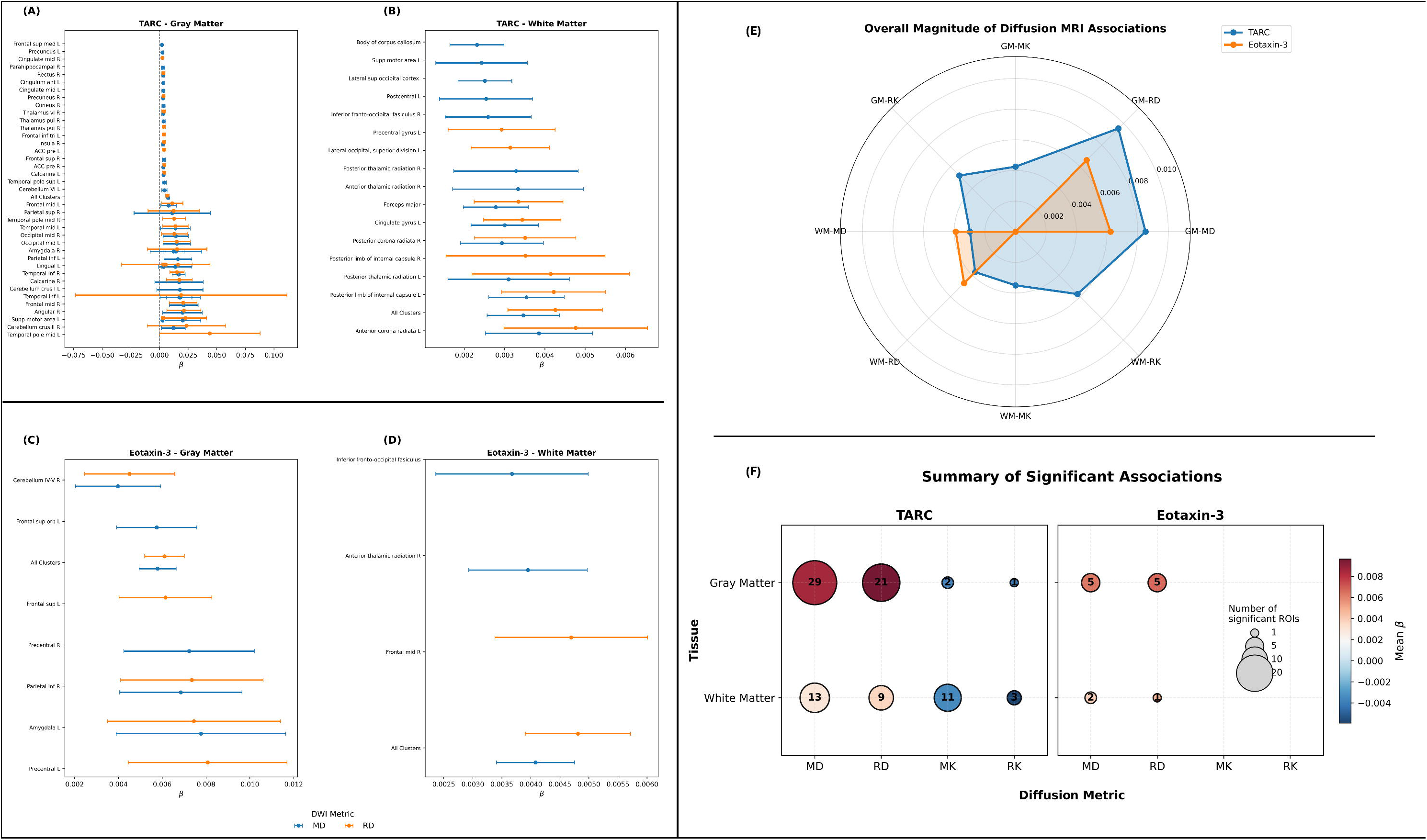
Cluster-based associations between systemic inflammatory chemokines and diffusion MRI metrics. (A) and (B) Forest plots showing significant cluster-level associations between circulating **TARC (CCL17)** levels and diffusion MRI metrics, and (C) and (D) corresponding associations for **Eotaxin-3 (CCL26)**. Points represent standardized regression coefficients (β), and horizontal bars indicate 95% confidence intervals derived from linear regression models adjusted for age, sex, and education. **Mean diffusivity (MD; blue)** and **radial diffusivity (RD; orange)** are shown, with the dashed vertical line indicating no association (β = 0). Clusters were identified from voxel-wise analyses that survived **cluster-level family-wise error (FWE) correction (p < 0.05)** and are labeled according to their predominant anatomical locations. (E) Radar plot summarizing the overall spatial distribution of significant associations across diffusion MRI metrics for TARC and Eotaxin-3, illustrating the relative extent of affected white matter regions for each inflammatory marker. (F) Bubble plot summarizing the number and direction of significant associations across diffusion metrics. Bubble size represents the number of significant white matter regions, while bubble color represents the average standardized β coefficient, with warm colors indicating positive associations and cool colors indicating negative associations. Abbreviations: **MD**, mean diffusivity; **RD**, radial diffusivity; **MK**, mean kurtosis; **RK**, radial kurtosis; β, standardized regression coefficient; **FWE**, family-wise error correction.

#### 3.2.2 Eotaxin-3

Cluster-based analyses revealed more limited associations for Eotaxin-3 compared with TARC. Higher Eotaxin-3 levels were associated with increased MD and RD in a small number of gray matter regions, including frontal, parietal, and limbic areas, as well as in select white matter tracts such as the anterior thalamic radiation and inferior fronto-occipital fasciculus. Effect sizes were generally smaller and confidence intervals wider than those observed for TARC, indicating greater variability and less extensive microstructural involvement. Panels C and D in Figure 4 present the standardized regression coefficients and 95% confidence intervals for significant Eotaxin-3–diffusion metric associations identified in the voxel-wise analyses.

## 4 Discussion

This study examined the relationship between circulating inflammatory markers and brain microstructure in cognitively unimpaired older African Americans using a voxel-wise diffusion MRI framework followed by cluster-based ROI modeling. TARC and Eotaxin-3 showed the most consistent associations with diffusion metrics, with TARC demonstrating the strongest and most widespread effects. Many of the identified regions overlap with the default mode network, known to show early vulnerability in preclinical AD and to support episodic memory, suggesting that TARC-associated microstructural alterations may preferentially target networks at risk for early AD-related change (Brier et al., 2014; Racine et al., 2019).

These chemokines were dominantly positively associated with MD and RD across multiple cortical and subcortical regions, indicating reduced microstructural integrity and potential myelin vulnerability(Chu et al., 2022; Racine et al., 2019). These effects were confirmed by cluster-based analyses and were observed both in gray and white matter, suggesting a diffuse rather than regionally localized pathology, aligning with emerging models of early network-level vulnerability in preclinical neurodegenerative disease states (Jack et al., 2018). These associations are also consistent with prior DTI and other quantitative MRI studies in aging and AD, showing increased demyelination and reduced tissue integrity (Kantarci et al., 2014; Moallemian et al., 2026, 2023).

TARC (CCL17) is a chemokine involved in Th2 lymphocytes and regulatory T cell recruitment and plays a central role in immune cell trafficking and systemic inflammatory responses (Campbell et al., 1999; Imai et al., 1997). Although incompletely studied, elevated TARC levels have been reported in individuals with mild cognitive impairment and AD, and it has been included in blood-based biomarker panels for neurodegenerative disorders(Campanelli et al., 2025). Peripheral inflammatory mediators may contribute to neuroinflammation through blood–brain barrier interactions and immune-cell trafficking, while also disrupting oligodendrocyte function and myelin maintenance, processes that are critical for preserving white matter integrity and efficient neural communication(Banks, 2005; Franklin and ffrench-Constant, 2017; Heneka et al., 2015; Varatharaj and Galea, 2017). These mechanisms provide a biologically plausible explanation for the observed associations between higher TARC levels and increased diffusivity metrics.

Eotaxin-3/CCL26, which signals through CCR3 to recruit eosinophils and Th2 cells, shares the broader Th2 immune polarization context of TARC. Circulating eotaxins increase with aging and have been implicated in neurodegenerative processes (Huber et al., 2018). Although its associations were less spatially extensive than those of TARC, which may reflect differences in receptor distribution, signal abundance, or the regional specificity of CCR3-mediated neuroinflammatory processes in aging — both chemokines showed convergent relationships with diffusivity measures across overlapping frontal and limbic regions, suggesting a shared inflammatory pathway underlying early microstructural vulnerability for Th2-polarizing chemokines.

In contrast, kurtosis-based metrics showed fewer and more spatially selective associations, primarily involving TARC. Higher TARC levels were associated with lower radial kurtosis in WM regions, potentially reflecting subtle alterations in microstructural complexity, axonal organization, or myelin integrity (Fieremans et al., 2013).

Neither TARC nor Eotaxin-3 was associated with MoCA performance (TARC: β=-0.020 p=0.902; Eotaxin-3: β=-0.094 p=0.561), suggesting that inflammation-related microstructural alterations may precede detectable cognitive decline in this cognitively unimpaired cohort. Given evidence of elevated inflammatory marker levels among Black individuals across the lifespan(Wiley et al., 2025), these findings may reflect population-specific mechanisms of early brain vulnerability that merit investigation in larger and racially diverse cohorts. A major strength of this study is its focus on older African Americans, a population at elevated risk AD/ADRD risk that remains underrepresented in neuroimaging research(Barnes, 2022).

Several limitations should be acknowledged. The cross-sectional design limited causal inference, and the modest sample size (n=41) precludes statistical power for subgroup analyses despite correction for multiple comparisons. In addition, plasma cytokines provide indirect measures of neuroinflammation and may not fully capture central immune processes. We did not adjust for vascular or cardiometabolic risk factors (e.g., hypertension, diabetes) or white-matter-hyperintensity burden, which influence both systemic inflammation and diffusion metrics and may contribute to the observed associations. Because the cluster-based analyses extracted effect estimates from clusters defined as significant in the same voxel-wise models, these estimates are descriptive and non-independent rather than an independent validation. In addition, multiple inflammatory markers, diffusion metrics, and tissue compartments were examined, so the TARC and Eotaxin-3 findings should be regarded as hypothesis-generating pending replication in larger, independent cohorts. Finally, restricted variability in MoCA scores (range 19-29, mean 23.4, SD 3.1) likely reduced sensitivity to detect associations between inflammatory markers and cognition.

In summary, higher circulating TARC and Eotaxin-3 levels were associated with diffusion MRI markers of reduced microstructural integrity, particularly increased MD and RD, in the absence of detectable cognitive impairment. Suggesting reduced microstructural integrity and myelin vulnerability in relation to systemic Th2-polarizing inflammatory signaling. This combined voxel-wise and cluster-based framework provides a robust approach for linking peripheral immune activity to early brain changes relevant to aging and AD/ADRD risk.

## Conflict of Interest

The authors declare no conflicts of interest. MB is a J. Seward Johnson Postdoctoral Fellow in Aging Neuroscience. JRR is the Dianne Isakson Distinguished Professor.

## Data Availability Statement

The data and scripts supporting the findings of this study will be made available for research purposes upon reasonable request to the corresponding authors.

## Acknowledgement

The authors would like to thank Yoonhee Han and Martina Ishaq for their technical assistance and contributions to blood and plasma sample preparation. We are also grateful for the feedback and insights shared by the Aging & Brain Health Alliance Community Brain Health Educators and Outreach Team: Glenda Wright, Delores Hammonds, Jerome Perkins, Louches Powell, Reverend Glenn Wilson, and Catherine Willis. In addition, we are deeply grateful to the thousands of community members who have participated in our brain health events since 2006, and to the more than 450 community members who have enrolled to date as VIPs (Very Important Participants) in the Rutgers Aging & Brain Health Alliance study.

## Author Contribution

**Study design:** SM, JRR, MAG; **Data collection:** SM, MB, BAF, MAG; **Data analysis:** SM, SR, JRR, IMW; **Manuscript preparation and editing:** SM, SR, IMW, MB, BAF, JRR, MAG.

## Funding

This study was supported by the National Institutes of Health (NIH) and the National Institute on Aging (NIA), under grant number R01AG053961 (NIH/NIA). Additional support was provided by R01NS130713, R01AG091689, R01AG0852729, R13AG069380, and the Dianne Isakson Distinguished Professorship.

**Preprint Reference: medRxiv; DOI:** https://doi.org/10.64898/2026.05.12.26353011

## Notes

### Competing Interest Statement

The authors have declared no competing interest.

### Author Declarations

The study was approved by the Rutgers Institutional Review Board (eIRB) and conducted in accordance with the principles of the 1975 Helsinki Declaration.

### Summary of Updates

This version of the manuscript contains updated analysis and results along with updated figures and tables.

