## Appendix for "Peripheral TARC (CCL17) Levels Are Associated with Widespread Microstructural Vulnerability in Cognitively Unimpaired Older African Americans"

**Supplementary Materials**

**Appendix**

### 1. Methods and Materials

#### 1.1. Participants

Inclusion criteria were: (1) Age ≥ 60 years, (2) availability of diffusion MRI and plasma biomarker data, (3) English fluency, 4) cognitively unimpaired status defined using education-adjusted Montreal Cognitive Assessment (MoCA) cutoffs validated for non-Hispanic Black individuals ^1^.

Exclusion criteria included use of cognitive-enhancing medications, history of learning disability, significant substance abuse, or recent exposure to general anesthesia (<3 months). Forty-one cognitively unimpaired older African American participants (29 female) met the inclusion criteria.

#### 1.2. MRI Acquisition and Curation

The MRI data were acquired using a 3T whole-body Prisma running VE11C scanner equipped with a standard 32-channel head coil at the Rutgers University Brain Imaging Center (RUBIC), in Newark. Whole-brain acquisitions included a T1-weighted (T1w) magnetization-prepared rapid acquisition gradient echo (MP-RAGE) sequence ^2^, with parameters: voxel size = 1 × 1 × 1 mm³, FOV = 240 × 256 × 208 mm³, TR = 2300 ms, TE = 2.98 ms, and TI = 900 ms. The T1-weighted acquisition was followed by a whole-brain diffusion-weighted imaging (DWI) scan, employing simultaneous multi-slice (SMS) acceleration and adaptive coil combination. The diffusion scan was performed in the transverse orientation with a field of view of 232 mm in both X and Y directions, and 162 mm in the Z direction, covering 81 contiguous slices with 2.0 mm isotropic resolution. The acquisition used echo and repetition times of 71 ms and 3400 ms, respectively. A total of 133 volumes were acquired, including 7 non-diffusion-weighted (b = 0 s/mm²) images and 126 diffusion-weighted volumes distributed across three b-value shells: 6 directions at b = 500 s/mm², 48 directions at b = 1000 s/mm², and 60 directions at b = 2000 s/mm². Diffusion directions were evenly distributed across shells using an electrostatic repulsion model and were temporally interleaved to minimize gradient heating. The scan employed a slice acceleration factor of 3 (SMS=3), with no in-plane acceleration (GRAPPA off), and utilized strong fat saturation and standard pre-scan normalization settings per the ADNI-4 protocol for the Alzheimer's Disease Neuroimaging Initiative.

Collected MRI data are anonymized and organized according to the Brain Imaging Data Structure (BIDS) ^3^ format using the DCM2BIDS tool (<https://unfmontreal.github.io/Dcm2Bids/3.2.0/> ) and an in-house developed pipeline available on GitHub (<https://github.com/Aging-and-Brain-Health-Alliance/Bidsification> ). Data is available from the corresponding authors upon request.

##### 1.2.1. Tissue Mask Generation

To examine tissue specific effects. Gray matter (GM) and white matter (WM) masks were derived from the tissue probability maps provided in SPM12. The GM and WM probability maps were thresholded and binarized to generate tissue-specific masks. The GM mask was defined using a probability threshold of 0.3, whereas the WM mask was defined using a more conservative threshold of 0.7, balancing sensitivity and anatomical specificity while minimizing partial volume effects at tissue boundaries. Because diffusion-derived maps were computed at 2 mm isotropic resolution, both masks were resampled to match the spatial resolution and voxel grid of the diffusion data using nearest-neighbor interpolation and were visually inspected to ensure appropriate anatomical coverage and alignment.

#### 1.3. Result Visualization

Significant clusters surviving the voxel-wise threshold (p < 0.001, uncorrected) and cluster-level family-wise error (FWE) correction (p < 0.05) were overlaid on the MNI anatomical template image to illustrate the spatial distribution of associations between inflammatory markers and diffusion metrics.

Cluster-Based results were visualized using forest plots. For each significant cluster, regression-derived effect estimates and corresponding confidence intervals were plotted, allowing direct comparison of the magnitude and direction of associations across diffusion metrics, tissue compartments (GM and WM), and inflammatory markers (TARC and Eotaxin-3).

##### 1.3.1. Diffusion Preprocessing

Diffusion data were preprocessed using QSIPrep (version 1.1.1)^4^, a robust and reproducible pipeline, which is built on Nipype (version 1.9.1) and follows the NiPreps framework. The pipeline integrates tools from FSL, MRtrix3, and ANTs within a standardized framework.

Briefly, diffusion images were denoised using the Marchenko-Pastur PCA approach^5,6^, followed by gibbs ringing correction. Bias field inhomogeneity was corrected using N4 algorithm^7^ as implemented in MRtrix3.

Susceptibility-included distortions were estimated using TOPUP based on pairs of images acquired with reversed phase-encoding directions. Head motion and eddy-current distortions were corrected using eddy^8,9^, including outlier detection and replacement. All corrections were performed within a unified framework, and the resulting data were resampled to AC-PC space with 2 mm isotropic resolution.

Confounding time series, including framewise displacement, motion parameters, and slice-wise cross-correlation metrics, were estimated and retained for quality control. Additional internal operations within QSIPrep utilized Nilearn^10^ and DIPY^11^.

### Results

#### 1.4. Whole Brain Voxel-based Analyses

##### 1.4.1. Mean Diffusivity and TARC

Table A-1. Summary statistics from the univariate GLM analysis between mean diffusivity and TARC within GM. The table shows the positive associations after p< 0.05 correction for family-wise error rate at the cluster-level. AAL3 atlas ^12^ is used to report GM regions.

| **Cluster p** | **k** | **Peak p** | **Peak T** | **X(mm)** | **Y(mm)** | **Z(mm)** | **Brain Region** |
| --- | --- | --- | --- | --- | --- | --- | --- |
| 0 | 190 | 0 | 9.7 | -35 | -84 | 31 | Occipital mid L |
| 0 | 194 | 0 | 9.54 | 13 | 40 | 3 | ACC_pre_R |
| 0 | 133 | 0 | 9.49 | -9 | 36 | -3 | Cingulum ant L |
| 0 | 230 | 0 | 9.33 | 53 | 0 | -37 | Temporal_Inf_R |
| 0 | 148 | 0 | 9.11 | -13 | -82 | -45 | Calcarine_R |
| 0 | 189 | 0 | 9.1 | 9 | -86 | -37 | Cerebelum_Crus2_R |
| 0 | 128 | 0 | 9.1 | -11 | 20 | 65 | Supp_Motor_Area_L |
| 0 | 97 | 0 | 9.08 | -49 | -16 | -37 | Amygdala_R |
| 0 | 40 | 0 | 8.9 | -47 | 6 | -41 | Temporal_Inf_L |
| 0 | 205 | 0 | 8.62 | 27 | -96 | -13 | Occipital_Mid_R |
| 0 | 147 | 0 | 8.41 | -17 | -28 | 5 | Thal_PuI_R |
| 0 | 101 | 0 | 8.31 | 43 | -64 | 51 | Angular_R |
| 0 | 357 | 0 | 8.27 | -13 | -96 | -15 | Lingual L |
| 0 | 82 | 0 | 8.12 | -41 | -34 | -27 | Temporal_Inf_L |
| 0.032 | 19 | 0 | 8.04 | 3 | 48 | 7 | Temporal_Pole_Sup_L |
| 0 | 48 | 0 | 7.99 | 15 | -50 | 39 | Precuneus_R |
| 0.006 | 25 | 0 | 7.94 | -35 | -32 | -13 | Insula_R |
| 0 | 91 | 0 | 7.82 | -7 | -24 | 47 | Cingulate_Mid_L |
| 0 | 43 | 0 | 7.62 | 11 | -16 | 37 | Frontal_Sup_Medial_L |
| 0 | 507 | 0 | 7.53 | 41 | 18 | 53 | Frontal_Mid_2_R |
| 0 | 54 | 0.001 | 7.45 | 3 | 38 | -17 | Rectus_R |
| 0 | 39 | 0.001 | 7.36 | -59 | 4 | -27 | Temporal_Mid_L |
| 0.013 | 22 | 0.001 | 7.32 | 33 | -38 | -9 | ParaHippocampal_R |
| 0 | 42 | 0.001 | 7.25 | -51 | -66 | -31 | Cerebelum_Crus1_L |
| 0.042 | 18 | 0.003 | 6.96 | -37 | -48 | -27 | Cerebelum_6_L |
| 0 | 228 | 0.003 | 6.95 | -19 | -68 | 63 | Parietal_Inf_L |
| 0 | 143 | 0.003 | 6.95 | 29 | -60 | 65 | Parietal_Sup_R |
| 0.024 | 20 | 0.003 | 6.95 | -29 | 32 | 51 | Frontal_Mid_2_L |
| 0 | 114 | 0.003 | 6.94 | 37 | 12 | -45 | Amygdala_R |
| 0 | 35 | 0.022 | 6.27 | -11 | 0 | 47 | Supp_Motor_Area_L |
| 0 | 59 | 0.043 | 6.06 | -11 | -80 | -3 | Lingual_L |
| 0 | 100 | 0.15 | 5.65 | 13 | -16 | 13 | Thal_VL_R |
| 0 | 40 | 0.184 | 5.58 | -17 | -68 | 17 | Calcarine_L |
| 0.008 | 24 | 0.198 | 5.56 | -7 | -60 | 47 | Precuneus_L |
| 0.032 | 19 | 0.82 | 5.1 | 11 | -62 | 9 | Cuneus_R |
| 0.032 | 19 | 1 | 4.67 | -25 | -2 | 49 | Frontal_Sup_2_R |
| **Key: mid: middle; ACC: anterior cingulate cortex; ant: anterior; inf: inferior; supp: supplementary; pul: Pulvinar lateral; sup: superior; L: left; R: right** | | | | | | | |

Table A-2. Summary statistics from the univariate GLM analysis between mean diffusivity and TARC within WM. The table shows the positive associations after p< 0.05 correction for family-wise error rate at the cluster-level. JHU WM tractography atlas ^13^ is used to report WM tracts.

| **Cluster p** | **k** | **Peak p** | **Peak T** | **x(mm)** | **y(mm)** | **z(mm)** | **Brain Region** |
| --- | --- | --- | --- | --- | --- | --- | --- |
| 0 | 739 | 0 | 9.28 | -13 | 38 | -5 | Forceps minor |
| 0 | 310 | 0 | 9.2 | -19 | -16 | 15 | Anterior thalamic radiation L |
| 0 | 97 | 0 | 9.09 | -17 | -44 | 39 | Cingulate gyrus L |
| 0.001 | 30 | 0 | 7.48 | -35 | -70 | 29 | Superior lateral occipital L |
| 0.005 | 22 | 0 | 7.47 | -11 | -10 | 35 | body of corpus callosum |
| 0.021 | 17 | 0 | 7.2 | 17 | -82 | 13 | Forceps major |
| 0 | 38 | 0.001 | 7 | 17 | -46 | 37 | Cingulate gyrus R |
| 0.028 | 16 | 0.014 | 6.03 | -19 | -46 | 49 | Postcentral gyrus L |
| 0.038 | 15 | 0.049 | 5.62 | 31 | 36 | -1 | Inferior fronto-occipital fasciculus R |
| 0.002 | 26 | 0.08 | 5.46 | 17 | -12 | 17 | Anterior thalamic radiation R |
| 0.003 | 24 | 0.137 | 5.29 | -11 | -16 | 49 | Superior corona radiata L |
| 0.007 | 21 | 0.16 | 5.24 | 31 | -58 | 11 | Inferior fronto-occipital fasciculus R |
| 0 | 38 | 0.213 | 5.14 | -33 | -62 | 5 | Inferior longitudinal fasciculus L/Inferior fronto-occipital fasciculus L/Forceps major |

##### 1.4.2. Radial Diffusivity and TARC

Table A-3. Summary statistics from the univariate GLM analysis between radial diffusivity and TARC within GM. The table shows the positive associations after p< 0.05 correction for family-wise error rate at the cluster-level. AAL3 atlas ^12^ is used to report GM regions.

| **Cluster p** | **k** | **Peak p** | **Peak T** | **x(mm)** | **y(mm)** | **z(mm)** | **Brain Region** |
| --- | --- | --- | --- | --- | --- | --- | --- |
| 0 | 191 | 0 | 10.42 | -35 | -84 | 31 | Occipital_Mid_L |
| 0 | 348 | 0 | 9.5 | -11 | -96 | -17 | Lingual_L |
| 0 | 187 | 0 | 9.11 | 17 | -84 | -43 | Cerebelum_Crus2_R |
| 0 | 246 | 0 | 8.96 | 53 | -6 | -39 | Temporal_Inf_R |
| 0 | 80 | 0 | 8.95 | -41 | -34 | -27 | Temporal_Inf_L |
| 0 | 129 | 0 | 8.9 | -11 | 20 | 65 | Supp_Motor_Area_L |
| 0 | 98 | 0 | 8.85 | -51 | -14 | -39 | Amygdala_R |
| 0 | 208 | 0 | 8.64 | 27 | -96 | -13 | Occipital_Mid_R |
| 0 | 157 | 0 | 8.52 | 11 | 38 | 7 | ACC_pre_R |
| 0 | 192 | 0 | 8.43 | -13 | -82 | -45 | Calcarine_R |
| 0 | 124 | 0 | 8.4 | 43 | -64 | 51 | Angular_R |
| 0 | 107 | 0 | 8.29 | -11 | 40 | 5 | ACC_pre_L |
| 0 | 140 | 0 | 8.17 | -17 | -28 | 5 | Thal_PuI_R |
| 0 | 94 | 0 | 7.77 | -15 | -20 | 39 | Frontal_Inf_Tri_L |
| 0 | 40 | 0 | 7.6 | -59 | 4 | -27 | Temporal_Mid_L |
| 0 | 543 | 0 | 7.59 | 47 | 20 | 47 | Frontal_Mid_2_R |
| 0 | 53 | 0 | 7.55 | 3 | 38 | -17 | Rectus_R |
| 0.017 | 22 | 0 | 7.55 | -35 | -32 | -13 | Insula_R |
| 0 | 38 | 0.001 | 7.36 | -51 | 12 | -35 | Temporal_Pole_Mid_L |
| 0 | 104 | 0.001 | 7.28 | 41 | 22 | -35 | Temporal_Pole_Mid_R |
| 0 | 39 | 0.003 | 6.96 | 15 | -50 | 39 | Precuneus_R |
| 0 | 71 | 0.003 | 6.95 | -39 | 18 | 53 | Frontal_Mid_2_L |
| 0 | 229 | 0.003 | 6.95 | -13 | -68 | 65 | Lingual_L |
| 0 | 152 | 0.003 | 6.95 | 33 | -54 | 65 | Parietal_Sup_R |
| 0 | 43 | 0.014 | 6.41 | 11 | -16 | 47 | Cingulate_Mid_R |
| 0 | 51 | 0.082 | 5.84 | -11 | -80 | -3 | Lingual_L |
| 0 | 93 | 0.107 | 5.76 | 13 | -16 | 13 | Thal_VL_R |
| 0.002 | 31 | 0.127 | 5.7 | -11 | 0 | 47 | Supp_Motor_Area_L |
| 0 | 37 | 0.167 | 5.61 | -5 | -78 | 25 | Calcarine_L |
| **Key: mid: middle; ACC: anterior cingulate cortex; ant: anterior; inf: inferior; supp: supplementary; pul: Pulvinar lateral; sup: superior; L: left; R: right** | | | | | | | |

Table A-4. Summary statistics from the univariate GLM analysis between radial diffusivity and TARC within WM. The table shows the positive associations after p< 0.05 correction for family-wise error rate at the cluster-level. JHU WM tractography atlas ^13^ is used to report WM tracts.

| **Cluster p** | **k** | **Peak p** | **Peak T** | **x(mm)** | **y(mm)** | **z(mm)** | **Brain Region** |
| --- | --- | --- | --- | --- | --- | --- | --- |
| 0 | 298 | 0 | 8.65 | -17 | -12 | 15 | Anterior thalamic radiation L |
| 0 | 127 | 0 | 8.34 | -17 | -56 | 39 | Cingulate gyrus L |
| 0 | 685 | 0 | 8.09 | -17 | 40 | -1 | Forceps minor/Uncinate fasiculus L/Inferior fronto-occipital fasiculus L |
| 0.002 | 29 | 0 | 7.47 | -35 | -70 | 29 | unclassified |
| 0.048 | 16 | 0.001 | 7.09 | 17 | -82 | 13 | Forceps major |
| 0 | 40 | 0.001 | 6.92 | 17 | -46 | 37 | Cingulate gyrus R |
| 0.004 | 26 | 0.026 | 5.82 | -9 | -20 | 51 | Precentral gyrus L |
| 0.001 | 34 | 0.185 | 5.19 | -33 | -62 | 5 | Inferior longitudinal fasiculus L/Inferior fronto-occipital fasiculus L/Forceps major |
| 0.008 | 23 | 0.923 | 4.54 | 19 | -18 | 15 | Corticospinal tract R |

##### 1.4.3. Mean Diffusivity and Eotaxin-3

Table A-5. Summary statistics from the univariate GLM analysis between mean diffusivity and Eotaxin-3 within GM. The table shows the positive associations after p< 0.05 correction for family-wise error rate at the cluster-level. AAL3 atlas ^12^ is used to report GM regions.

| **Cluster p** | **k** | **Peak p** | **Peak T** | **x(mm)** | **y(mm)** | **z(mm)** | **Brain Region** |
| --- | --- | --- | --- | --- | --- | --- | --- |
| 0.033 | 19 | 0.001 | 7.25 | 35 | -14 | 43 | Precentral R |
| 0.024 | 20 | 0.027 | 6.21 | -29 | 4 | -21 | Amygdala L |
| 0 | 71 | 0.249 | 5.48 | 9 | -54 | -21 | Cerebellum IV-V R |
| 0.011 | 23 | 0.354 | 5.37 | -25 | 54 | -3 | Frontal sup orb L |
| 0.033 | 19 | 0.973 | 4.97 | 37 | -44 | 47 | Parietal inf R |
|  |  | 0.482 | 5.27 | -3 | -54 | -19 | Vermis IV-V |

Table A-6. Summary statistics from the univariate GLM analysis between mean diffusivity and Eotaxin-3 within WM. The table shows the positive associations after p< 0.05 correction for family-wise error rate at the cluster-level. JHU WM tractography atlas ^13^ is used to report WM tracts.

| **Cluster p** | **k** | **Peak p** | **Peak T** | **x(mm)** | **y(mm)** | **z(mm)** | **Brain Region** |
| --- | --- | --- | --- | --- | --- | --- | --- |
| 0.009 | 20 | 0 | 7.55 | 27 | 30 | 25 | Anterior thalamic radiation R |
| 0.04 | 15 | 0.045 | 5.65 | 31 | 48 | -3 | Anterior thalamic radiation R/Inferior fronto-occipital fasciculus |

##### 1.4.4. Radial Diffusivity and Eotaxin-3

Table A-7. Summary statistics from the univariate GLM analysis between radial diffusivity and Eotaxin-3 within GM. The table shows the positive associations after p< 0.05 correction for family-wise error rate at the cluster-level. AAL3 atlas ^12^ is used to report GM regions.

| **Cluster p** | **k** | **Peak p** | **Peak T** | **x(mm)** | **y(mm)** | **z(mm)** | **Brain Region** |
| --- | --- | --- | --- | --- | --- | --- | --- |
| 0.041 | 19 | 0.003 | 6.88 | 35 | -14 | 43 | Precentral L |
| 0.01 | 24 | 0.105 | 5.77 | -29 | 2 | -19 | Amygdala L |
| 0 | 61 | 0.16 | 5.63 | 9 | -54 | -21 | Cerebelum_4_5_R |
| 0.008 | 25 | 0.603 | 5.2 | -25 | 54 | -3 | Frontal_Sup_2_L |
| 0.031 | 20 | 0.977 | 4.94 | 37 | -44 | 47 | Parietal_Inf_R |
| **Key: inf: inferior; sup: superior; L: left; R: right** | | | | | | | |

##### 1.4.5. Mean Kurtosis and TARC

Table A-8. Summary statistics from the univariate GLM analysis between mean kurtosis and TARC within GM. The table shows the positive associations after p< 0.05 correction for family-wise error rate at the cluster-level. AAL3 atlas ^12^ is used to report GM regions.

| **Cluster p** | **k** | **Peak p** | **Peak T** | **x(mm)** | **y(mm)** | **z(mm)** | **Brain Region** |
| --- | --- | --- | --- | --- | --- | --- | --- |
| 0.003 | 15 | 0.027 | 6.21 | -13 | -24 | 45 | Precentral_L |
| 0.01 | 13 | 1 | 4.85 | 13 | -62 | 13 | Precentral_R |

Table A-9. Summary statistics from the univariate GLM analysis between mean kurtosis and TARC within WM. The table shows the positive associations after p< 0.05 correction for family-wise error rate at the cluster-level. JHU WM tractography atlas ^13^ is used to report WM tracts.

| **Cluster p** | **k** | **Peak p** | **Peak T** | **x(mm)** | **y(mm)** | **z(mm)** | **Brain Region** |
| --- | --- | --- | --- | --- | --- | --- | --- |
| 0 | 30 | 0 | 7.79 | 19 | -12 | 15 | Anterior thalamic radiation R |
| 0.002 | 20 | 0 | 7.24 | -17 | -30 | 33 | Body of corpus callosum |
| 0 | 27 | 0 | 7.22 | 19 | -36 | 33 | Posterior corona radiata R |
| 0.007 | 16 | 0.001 | 6.85 | -23 | -18 | 17 | Corticospinal tract L |
| 0 | 38 | 0.051 | 5.61 | -17 | 38 | 9 | Forceps minor/ Cingulate gyrus L |
| 0 | 29 | 0.052 | 5.61 | -17 | -22 | 39 | Superior corona radiata L |
| 0.01 | 15 | 0.27 | 5.07 | -15 | -56 | 37 | Cingulate gyrus L |
| 0.021 | 13 | 0.302 | 5.03 | -9 | -78 | 21 | Cuneal cortex/intra calcarine cortex |
| 0 | 47 | 0.415 | 4.93 | 15 | 20 | 21 | Genu of corpus callosum |
| 0.01 | 15 | 0.538 | 4.84 | 17 | 42 | 1 | Forceps minor |
| 0.032 | 12 | 0.953 | 4.64 | 21 | 0 | 19 | Superior fronto-occipital fasciculus (could be a part of anterior internal capsule) R |

##### 1.4.6. Radial Kurtosis and TARC

Table A-10. Summary statistics from the univariate GLM analysis between radial kurtosis and TARC within WM. The table shows the positive associations after p< 0.05 correction for family-wise error rate at the cluster-level. JHU WM tractography atlas ^13^ is used to report WM tracts

| **Cluster p** | **k** | **Peak p** | **Peak T** | **x(mm)** | **y(mm)** | **z(mm)** | **Brain Region** |
| --- | --- | --- | --- | --- | --- | --- | --- |
| 0.009 | 14 | 0.822 | 4.7 | 15 | 20 | 21 | Genu of corpus callosum |
| 0.033 | 11 | 0.925 | 4.66 | 25 | -18 | 19 | Corticospinal tract R |
| 0.021 | 12 | 0.999 | 4.33 | -19 | -36 | 35 | Posterior corona radiata L |

##### 1.4.7. Other Inflammatory Markers

Among the remaining inflammatory markers examined, Tumor Necrosis Factor alpha (TNF-α), interleukin 1 beta (IL-1β), interleukin 10 (IL-10), and Monocyte Chemoattractant Protein 1 (MCP-1) did not show significant associations with any diffusion -derived metrics within GM and WM.

In contrast interleukin5 (IL-5) demonstrated significant positive associations with MD, RD, and MK both in WM and GM. Within GM, higher IL-5 levels were associated with increased MD and RD in the right middle occipital gyrus and left insula. In WM, significant associations were consistently observed within the left superior longitudinal fasciculus. Similarly, MK showed localized positive associations with IL-5 within the left insula in GM and within the left superior longitudinal fasciculus in WM.

Table A-11. Summary statistics from the univariate GLM analysis between mean diffusivity and IL-5 within GM and WM. The table shows the positive associations after p< 0.05 correction for family-wise error rate at the cluster-level. AAL3 atlas ^12^ is used to report GM regions. JHU WM tractography atlas ^13^ is used to report WM tracts

| **Cluster p** | **k** | **Peak p** | **Peak T** | **x(mm)** | **y(mm)** | **z(mm)** | **Brain Region** |
| --- | --- | --- | --- | --- | --- | --- | --- |
| **Positive association between MD and IL-5 within GM** | | | | | | | |
| 0 | 51 | 0.181 | 5.61 | 41 | -76 | 3 | Middle Occipital Gyrus R |
| 0.001 | 31 | 0.191 | 5.16 | -39 | 2 | 7 | Insula L |
| **Positive association between MD and IL-5 within WM** | | | | | | | |
| 0.009 | 20 | 0.0005 | 6.38 | -41 | -42 | 27 | Superior longitudinal fasciculus L |

Table A-12. Summary statistics from the univariate GLM analysis between radial diffusivity and IL-5 within GM and WM. The table shows the positive associations after p< 0.05 correction for family-wise error rate at the cluster-level. AAL3 atlas ^12^ is used to report GM regions. JHU WM tractography atlas ^13^ is used to report WM tracts

| **Cluster p** | **k** | **Peak p** | **Peak T** | **x(mm)** | **y(mm)** | **z(mm)** | **Brain Region** |
| --- | --- | --- | --- | --- | --- | --- | --- |
| **Positive association between RD and IL-5 within GM** | | | | | | | |
| 0.002 | 31 | 0.278 | 5.25 | -41 | 4 | 7 | Insula L |
| 0 | 43 | 0.278 | 5.16 | 41 | -74 | 3 | Inferior Occipital Gyrus R |
| **Positive association between RD and IL-5 within WM** | | | | | | | |
| 0.011 | 22 | 0.013 | 6.06 | -41 | -44 | 27 | Superior longitudinal fasciculus L |

Table A-13. Summary statistics from the univariate GLM analysis between mean kurtosis and IL-5 within GM and WM. The table shows the positive associations after p< 0.05 correction for family-wise error rate at the cluster-level. AAL3 atlas ^12^ is used to report GM regions. JHU WM tractography atlas ^13^ is used to report WM tracts

| **Cluster p** | **k** | **Peak p** | **Peak T** | **x(mm)** | **y(mm)** | **z(mm)** | **Brain Region** |
| --- | --- | --- | --- | --- | --- | --- | --- |
| **Positive association between MK and IL-5 within GM** | | | | | | | |
| 0.017 | 12 | 0.116 | 5.73 | -55 | -30 | 35 | Insula L |
| **Positive association between MK and IL-5 within WM** | | | | | | | |
| 0.01 | 15 | 0.02 | 5.9 | -37 | -42 | 27 | Superior longitudinal fasciculus L |

As presented in Table A-14, Interleukin 6 (IL-6) showed significant associations with both RD and Md within small clusters bilaterally covering part of middle frontal regions.

Table A-14. Summary statistics from the univariate GLM analysis between mean and radial diffusivity and IL-6 within GM and WM. The table shows the positive associations after p< 0.05 correction for family-wise error rate at the cluster-level. AAL3 atlas ^12^ is used to report GM regions.

| **Cluster p** | **k** | **Peak p** | **Peak T** | **x(mm)** | **y(mm)** | **z(mm)** | **Brain Region** |
| --- | --- | --- | --- | --- | --- | --- | --- |
| **Positive association between MD and IL-6 within GM** | | | | | | | |
| 0.003 | 28 | 0.009 | 6.58 | -33 | 56 | 21 | Middle Frontal L |
| 0.001 | 33 | 0.398 | 5.33 | 47 | -78 | 19 | Middle Frontal R |
| **Positive association between RD and IL-6 within GM** | | | | | | | |
| 0.006 | 26 | 0.006 | 6.71 | -33 | 56 | 21 | Middle Frontal L |
| 0.001 | 35 | 0.475 | 5.27 | 43 | -84 | 17 | Middle Frontal R |

Increased levels of interleukin 7 (IL-7) were positively associated with higher MD and RD within multiple GM regions, including the temporal pole, inferior occipital gyrus, middle frontal cortex, and hippocampus. (Table A-15)

Table A-15. Summary statistics from the univariate GLM analysis between mean and radial diffusivity and IL-7 within GM and WM. The table shows the positive associations after p< 0.05 correction for family-wise error rate at the cluster-level. AAL3 atlas ^12^ is used to report GM regions.

| **Cluster p** | **k** | **Peak p** | **Peak T** | **x(mm)** | **y(mm)** | **z(mm)** | **Brain Region** |
| --- | --- | --- | --- | --- | --- | --- | --- |
| **Positive association between MD and IL-7 within GM** | | | | | | | |
| 0 | 154 | 0.026 | 6.22 | -51 | 8 | -37 | Temporal pole L |
| 0.018 | 21 | 0.17 | 5.61 | 29 | -94 | -11 | Inferior Occipital Gyrus R |
| 0 | 86 | 0.25 | 5.48 | 19 | -84 | -41 | Cerebellum Crus 2 R |
| 0 | 54 | 0.315 | 5.41 | -29 | -88 | -23 | Cerebellum Crus 1 L |
| 0 | 72 | 0.951 | 5.05 | -35 | -80 | -39 | Cerebellum Crus 2 L |
| 0 | 50 | 1 | 4.6 | 29 | 40 | 41 | Middle Frontal R |
| 0.024 | 20 | 1 | 4.34 | 19 | -32 | -5 | Temporal pole L |
| **Positive association between RD and IL-7 within GM** | | | | | | | |
| 0 | 141 | 0.002 | 6.99 | -51 | 8 | -37 | Temporal pole L |
| 0 | 73 | 0.198 | 5.56 | 19 | -84 | -41 | Cerebellum Crus 2 R |
| 0 | 52 | 0.378 | 5.35 | -29 | -88 | -23 | Cerebellum Crus 1 L |
| 0.006 | 26 | 0.812 | 5.1 | 11 | -58 | -59 | Cerebellum Lobule 8 R |
| 0 | 63 | 0.836 | 5.09 | -35 | -80 | -39 | Cerebellum Crus 2 L |
| 0 | 43 | 1 | 4.56 | 35 | 28 | 47 | Middle Frontal R |
| 0.039 | 19 | 1 | 4.24 | 19 | -32 | -5 | Hippocampus R |

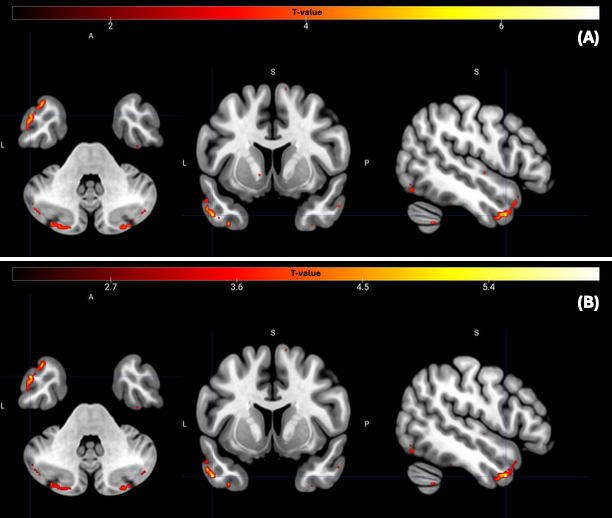

Figure A-1. **MD and RD signal increase with higher IL-7 levels in gray matter.** Statistical parametric maps identify regions within (A) MD and (B) RD where diffusivity metrics increase with higher IL-7 levels (p_uncor_<0 .001). T-value maps are overlaid on the MNI152 template. The color bar represents t-scores corresponding to the association with IL-7. Abbreviations: MD, mean diffusivity; RD, radial diffusivity; uncorr, uncorrected.

In addition, Macrophage Inflammatory Protein 1 alpha demonstrated widespread positive associations with MD across both gray matter and white matter. Within GM, higher MIP-1α levels were associated with increased MD in multiple cortical and subcortical regions, including the putamen, superior temporal gyrus, parahippocampal gyrus, anterior cingulate cortex, fusiform gyrus, temporal pole, and hippocampus. In WM, significant associations were observed across several major tracts, including the inferior fronto-occipital fasciculus, corticospinal tract, superior longitudinal fasciculus, anterior thalamic radiation, and posterior limb of the internal capsule. These effects were spatially extensive and characterized by high t-values across clusters, indicating a robust relationship between MIP-1α levels and increased diffusivity. (Table 16).

Table A-16. Summary statistics from the univariate GLM analysis between mean diffusivity and MIP- α within GM and WM. The table shows the positive associations after p< 0.05 correction for family-wise error rate at the cluster-level. AAL3 atlas ^12^ is used to report GM regions. JHU WM tractography atlas ^13^ is used to report WM tracts

| **Cluster p** | **k** | **Peak p** | **Peak T** | **x(mm)** | **y(mm)** | **z(mm)** | **Brain Region** |
| --- | --- | --- | --- | --- | --- | --- | --- |
| **Positive association between MD and MIP-1α within GM** | | | | | | | |
| 0 | 126 | 0 | 19.59 | 33 | -12 | -7 | Putamen R |
| 0.005 | 26 | 0 | 18.44 | -41 | -16 | -9 | Superior temporal L |
| 0.001 | 32 | 0 | 15.43 | 29 | -36 | -9 | Parahippocampal R |
| 0.001 | 32 | 0 | 14.34 | -7 | 28 | 19 | Anterior cingulate L |
| 0 | 69 | 0 | 11.76 | -41 | -54 | 11 | Superior temporal L |
| 0.004 | 27 | 0 | 11.47 | -37 | -12 | -43 | Fusiform L |
| 0 | 41 | 0 | 11.44 | -25 | -40 | -5 | Temporal pole L |
| 0 | 45 | 0 | 10.09 | 25 | -12 | -25 | Hippocampus R |
| 0.006 | 25 | 0 | 10.09 | 29 | -62 | -31 | Temporal pole R |
| **Positive association between MD and MIP-1α within WM** | | | | | | | |
| 0 | 89 | 0 | 33.46 | 29 | -26 | 5 | Inferior fronto-occipital fasciculus R |
| 0 | 33 | 0 | 17.61 | 27 | -14 | 17 | Corticospinal tract R |
| 0.029 | 16 | 0 | 14.69 | -41 | -54 | 9 | Superior longitudinal fasciculus L |
| 0.009 | 20 | 0 | 12.25 | 43 | 16 | 17 | Superior longitudinal fasciculus R |
| 0 | 38 | 0 | 11.54 | 7 | -30 | -15 | Anterior thalamic radiation R |
| 0.003 | 24 | 0 | 9.91 | 21 | -20 | 3 | Posterior limb of internal capsule R |
| 0.001 | 29 | 0 | 8.87 | -41 | -48 | 27 | Superior longitudinal fasciculus L |
| 0.001 | 28 | 0 | 7.51 | 37 | -42 | 25 | Superior longitudinal fasciculus R |

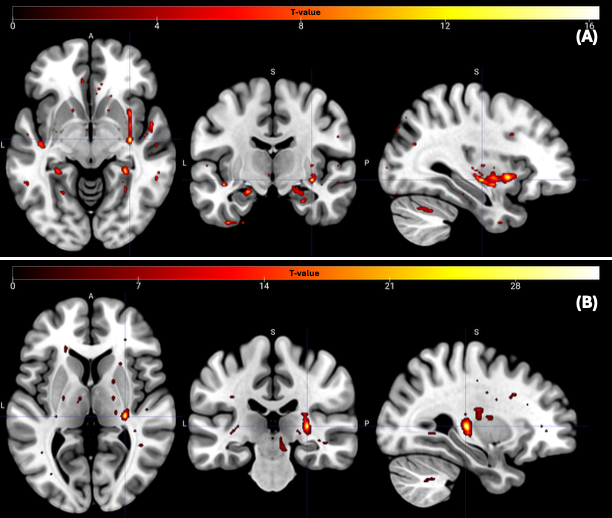

Figure A-2. **MD signal increase with higher MIP-1α levels in gray and white matter.** Statistical parametric maps identify regions within (A) gray matter and (B) white matter where diffusivity metrics increase with higher MIP-1α levels (p_uncor_<0 .001). T-value maps are overlaid on the MNI152 template. The color bar represents t-scores corresponding to the association with IL-7. Abbreviations: MD, mean diffusivity; uncorr, uncorrected.

#### 1.5. Cluster-Based Analyses

##### 1.5.1. TARC

Table A-17. Cluster-Based associations between TARC and mean diffusivity within gray matter. Mean diffusion values were extracted from significant clusters identified in voxel-wise analyses and entered into regression models controlling for age, sex, and education. β coefficients represent standardized effects. GM, gray matter; CI, confidence interval.

| **Region** | **Hemisphere** | **Beta** | **SE** | **CI low** | **CI high** | **P** |
| --- | --- | --- | --- | --- | --- | --- |
| **All Clusters** |  | 0.00756216 | 0.00055009 | 0.00644653 | 0.0086778 | 6.78E-16 |
| **ACC pre** | Right | 0.0033021 | 0.00041656 | 0.00245727 | 0.00414692 | 2.08E-09 |
| **Amygdala** | Right | 0.01437084 | 0.00808635 | -0.0080805 | 0.03682214 | 0.15017833 |
| **Amygdala** | Right | 0.01232162 | 0.00421259 | 0.00314318 | 0.02150006 | 0.01272273 |
| **Angular** | Right | 0.0201113 | 0.00674292 | 0.00277809 | 0.03744452 | 0.03070842 |
| **Calcarine** | Left | 0.00348523 | 0.00053016 | 0.00241001 | 0.00456045 | 1.20E-07 |
| **Calcarine** | Right | 0.01710185 | 0.00864851 | -0.0040603 | 0.03826399 | 0.09536248 |
| **Cerebellum VI** | Left | 0.00431084 | 0.00110805 | 0.00206361 | 0.00655807 | 0.00041451 |
| **Cerebellum crus I** | Left | 0.01783237 | 0.00785125 | -0.0023499 | 0.03801465 | 0.0723231 |
| **Cerebellum crus II** | Right | 0.01206628 | 0.00503005 | 0.0016346 | 0.02249795 | 0.02535922 |
| **Cingulate mid** | Left | 0.00338671 | 0.00060659 | 0.0021565 | 0.00461692 | 2.51E-06 |
| **Cingulum ant** | Left | 0.00336443 | 0.00046005 | 0.0024314 | 0.00429746 | 1.28E-08 |
| **Cuneus** | Right | 0.00346796 | 0.00067769 | 0.00209355 | 0.00484238 | 1.05E-05 |
| **Frontal mid** | Left | 0.00795658 | 0.00249896 | 0.00101834 | 0.01489481 | 0.03340877 |
| **Frontal mid** | Right | 0.02123851 | 0.00590966 | 0.00871059 | 0.03376644 | 0.00243053 |
| **Frontal sup med** | Left | 0.00209241 | 0.00028055 | 0.00152342 | 0.00266139 | 8.30E-09 |
| **Frontal sup** | Right | 0.00396546 | 0.00069287 | 0.00256026 | 0.00537067 | 1.63E-06 |
| **Insula** | Right | 0.00303797 | 0.00054012 | 0.00194256 | 0.00413337 | 2.21E-06 |
| **Lingual** | Left | 0.01372252 | 0.00690241 | -0.0007789 | 0.02822394 | 0.06223016 |
| **Lingual** | Left | 0.00330162 | 0.00062702 | 0.00202997 | 0.00457327 | 6.66E-06 |
| **Occipital mid** | Left | 0.01522311 | 0.00565012 | 0.00318016 | 0.02726605 | 0.01664802 |
| **Occipital mid** | Right | 0.01427879 | 0.00514148 | 0.00325141 | 0.02530617 | 0.01483157 |
| **Parahippocampal** | Right | 0.00289483 | 0.00059464 | 0.00168885 | 0.00410082 | 2.25E-05 |
| **Parietal inf** | Left | 0.01605648 | 0.00551211 | 0.00392441 | 0.02818856 | 0.01411443 |
| **Parietal sup** | Right | 0.01108965 | 0.01296595 | -0.0222404 | 0.04441967 | 0.4314558 |
| **Precuneus** | Left | 0.00255242 | 0.00057308 | 0.00139017 | 0.00371467 | 7.86E-05 |
| **Precuneus** | Right | 0.00310312 | 0.00043266 | 0.00222564 | 0.00398061 | 1.95E-08 |
| **Rectus** | Right | 0.00325141 | 0.00056808 | 0.00209928 | 0.00440354 | 1.63E-06 |
| **Supp motor area** | Left | 0.0204103 | 0.00689936 | 0.00480286 | 0.03601774 | 0.01600398 |
| **Supp motor area** | Left | 0.00255466 | 0.0005581 | 0.00142279 | 0.00368653 | 5.42E-05 |
| **Temporal inf** | Left | 0.01726589 | 0.00495704 | 0.00622093 | 0.02831086 | 0.00589049 |
| **Temporal inf** | Left | 0.01822074 | 0.00684724 | 0.00061936 | 0.03582211 | 0.04482814 |
| **Temporal inf** | Right | 0.01674487 | 0.0026588 | 0.01107777 | 0.02241197 | 1.43E-05 |
| **Temporal mid** | Left | 0.01391138 | 0.00567496 | 0.00082491 | 0.02699785 | 0.03985193 |
| **Temporal pole sup** | Left | 0.0041424 | 0.0008867 | 0.00234409 | 0.00594071 | 4.08E-05 |
| **Thalamus pul** | Right | 0.00357385 | 0.00067499 | 0.00220491 | 0.00494279 | 6.09E-06 |
| **Thalamus vl** | Right | 0.00331625 | 0.0007021 | 0.00189232 | 0.00474019 | 3.49E-05 |

Table A-18. Cluster-level associations between TARC and mean diffusivity within white matter. Mean diffusion values were extracted from significant clusters identified in voxel-wise analyses and entered into regression models controlling for age, sex, and education. β coefficients represent standardized effects. WM, white matter; MD, mean diffusivity.

| **Region** | **Hemisphere** | **Beta** | **SE** | **CI low** | **CI high** | **P** |
| --- | --- | --- | --- | --- | --- | --- |
| **All Clusters** |  | 0.00346659 | 0.0004441 | 0.00256592 | 0.00436726 | 2.97E-09 |
| **Anterior corona radiata** | Left | 0.00385375 | 0.00065511 | 0.00252513 | 0.00518237 | 9.97E-07 |
| **Anterior thalamic radiation** | Right | 0.00333533 | 0.00080184 | 0.00170912 | 0.00496154 | 0.00018873 |
| **Body of corpus callosum** |  | 0.00231346 | 0.00033084 | 0.00164248 | 0.00298443 | 3.36E-08 |
| **Cingulate gyrus** | Left | 0.00300501 | 0.00041336 | 0.00216668 | 0.00384333 | 1.46E-08 |
| **Forceps major** |  | 0.00278114 | 0.00039728 | 0.00197541 | 0.00358687 | 3.28E-08 |
| **Inferior fronto-occipital fasiculus** | Right | 0.0025935 | 0.00052661 | 0.00152548 | 0.00366153 | 1.89E-05 |
| **Lateral sup occipital cortex** |  | 0.00251148 | 0.00032925 | 0.00184373 | 0.00317922 | 5.01E-09 |
| **Postcentral** | Left | 0.00254207 | 0.00057058 | 0.00138487 | 0.00369926 | 7.83E-05 |
| **Posterior corona radiata** | Right | 0.00293244 | 0.00050813 | 0.00190192 | 0.00396297 | 1.41E-06 |
| **Posterior limb of internal capsule** | Left | 0.0035441 | 0.00046362 | 0.00260384 | 0.00448437 | 4.78E-09 |
| **Posterior thalamic radiation** | Left | 0.00310018 | 0.00074282 | 0.00159368 | 0.00460667 | 0.00018111 |
| **Posterior thalamic radiation** | Right | 0.00328297 | 0.00076268 | 0.00173618 | 0.00482975 | 0.00012283 |
| **Supp motor area** | Left | 0.00242874 | 0.00056062 | 0.00129174 | 0.00356574 | 0.00011312 |

Table A-19. Cluster-level associations between TARC and radial diffusivity within gray matter. Mean diffusion values were extracted from significant clusters identified in voxel-wise analyses and entered into regression models controlling for age, sex, and education. β coefficients represent standardized effects. GM, gray matter; RD, radial diffusivity.

| **Region** | **Hemisphere** | **Beta** | **SE** | **CI low** | **CI high** | **P** |
| --- | --- | --- | --- | --- | --- | --- |
| **All Clusters** |  | 0.00695751 | 0.00079158 | 0.0053521 | 0.00856291 | 1.73E-10 |
| **ACC pre** | Left | 0.00389512 | 0.00081439 | 0.00224346 | 0.00554678 | 2.91E-05 |
| **ACC pre** | Right | 0.0040744 | 0.00063029 | 0.0027961 | 0.00535269 | 1.67E-07 |
| **Amygdala** | Right | 0.01537195 | 0.00937089 | -0.0106458 | 0.0413897 | 0.176267 |
| **Angular** | Right | 0.02131032 | 0.00610021 | 0.00638363 | 0.036237 | 0.01293016 |
| **Calcarine** | Left | 0.00410169 | 0.00073002 | 0.00262114 | 0.00558224 | 2.25E-06 |
| **Calcarine** | Right | 0.01736972 | 0.00489373 | 0.00608475 | 0.02865469 | 0.00751623 |
| **Cerebellum crus II** | Right | 0.02357309 | 0.01631137 | -0.0106958 | 0.05784202 | 0.16558865 |
| **Cingulate mid** | Right | 0.00256926 | 0.00037288 | 0.00181302 | 0.0033255 | 4.57E-08 |
| **Frontal inf tri** | Left | 0.00373739 | 0.00065057 | 0.00241799 | 0.0050568 | 1.52E-06 |
| **Frontal mid** | Left | 0.01117635 | 0.0042115 | 0.00190689 | 0.0204458 | 0.02243498 |
| **Frontal mid** | Right | 0.02067661 | 0.00576103 | 0.0085219 | 0.03283132 | 0.00226204 |
| **Insula** | Right | 0.00378248 | 0.00074268 | 0.00227625 | 0.00528871 | 1.13E-05 |
| **Lingual** | Left | 0.00546785 | 0.01838168 | -0.0331506 | 0.04408634 | 0.76951959 |
| **Lingual** | Left | 0.0161752 | 0.00566356 | 0.0037098 | 0.0286406 | 0.01562765 |
| **Lingual** | Left | 0.00346677 | 0.00069059 | 0.0020662 | 0.00486735 | 1.41E-05 |
| **Occipital mid** | Left | 0.01502502 | 0.00558993 | 0.00311038 | 0.02693967 | 0.0168637 |
| **Occipital mid** | Right | 0.01298148 | 0.00528401 | 0.00171888 | 0.02424408 | 0.02668517 |
| **Parietal sup** | Right | 0.01236284 | 0.00914313 | -0.0100096 | 0.03473529 | 0.22506849 |
| **Precuneus** | Right | 0.00342445 | 0.00058323 | 0.00224161 | 0.00460729 | 1.03E-06 |
| **Rectus** | Right | 0.00333046 | 0.00062869 | 0.00205542 | 0.00460549 | 6.04E-06 |
| **Supp motor area** | Left | 0.02268261 | 0.00818776 | 0.00416061 | 0.0412046 | 0.0217426 |
| **Supp motor area** | Left | 0.00284131 | 0.0007336 | 0.0013535 | 0.00432911 | 0.00043585 |
| **Temporal inf** | Left | 0.01888514 | 0.03597795 | -0.0735991 | 0.11136939 | 0.62208019 |
| **Temporal inf** | Right | 0.01526478 | 0.00294931 | 0.00904229 | 0.02148728 | 7.60E-05 |
| **Temporal mid** | Left | 0.01385056 | 0.00494574 | 0.00266253 | 0.02503859 | 0.02069551 |
| **Temporal pole mid** | Left | 0.04394605 | 0.01973726 | -3.13E-05 | 0.0879234 | 0.05013457 |
| **Temporal pole mid** | Right | 0.01274893 | 0.00460503 | 0.00271544 | 0.02278243 | 0.01700973 |
| **Thalamus pui** | Right | 0.003693 | 0.00068431 | 0.00230516 | 0.00508084 | 4.45E-06 |
| **Thalamus vl** | Right | 0.00355898 | 0.0007772 | 0.00198275 | 0.00513521 | 5.39E-05 |

Table A-20. Cluster-level associations between TARC and radial diffusivity within white matter. Mean diffusion values were extracted from significant clusters identified in voxel-wise analyses and entered into regression models controlling for age, sex, and education. β coefficients represent standardized effects. WM, white matter; RD, radial diffusivity.

| **Region** | **Hemisphere** | **Beta** | **SE** | **CI_low** | **CI_high** | **P** |
| --- | --- | --- | --- | --- | --- | --- |
| **All Clusters** |  | 0.00426024 | 0.00057962 | 0.00308471 | 0.00543576 | 1.15E-08 |
| **Anterior corona radiata** | Left | 0.0047674 | 0.00087778 | 0.00298717 | 0.00654762 | 4.00E-06 |
| **Cingulate gyrus** | Left | 0.00343967 | 0.00047296 | 0.00248046 | 0.00439888 | 1.45E-08 |
| **Forceps major** |  | 0.00334602 | 0.00054368 | 0.0022434 | 0.00444865 | 4.32E-07 |
| **Lateral occipital, superior division** | Left | 0.00314326 | 0.00048097 | 0.0021678 | 0.00411871 | 1.35E-07 |
| **Posterior corona radiata** | Right | 0.00350845 | 0.00062214 | 0.0022467 | 0.0047702 | 2.11E-06 |
| **Posterior limb of internal capsule** | Left | 0.0042201 | 0.00063637 | 0.00292948 | 0.00551073 | 1.00E-07 |
| **Posterior limb of internal capsule** | Right | 0.00352001 | 0.00097398 | 0.0015447 | 0.00549533 | 0.00091393 |
| **Posterior thalamic radiation** | Left | 0.00414654 | 0.00096554 | 0.00218834 | 0.00610473 | 0.00012653 |
| **Precentral gyrus** | Left | 0.0029261 | 0.00065487 | 0.00159796 | 0.00425423 | 7.53E-05 |

Table A-21. Cluster-level associations between TARC and mean kurtosis within gray matter. Mean diffusion values were extracted from significant clusters identified in voxel-wise analyses and entered into regression models controlling for age, sex, and education. β coefficients represent standardized effects. GM, gray matter; MK, mean kurtosis.

| **Region** | **Hemisphere** | **Beta** | **SE** | **CI low** | **CI high** | **P** |
| --- | --- | --- | --- | --- | --- | --- |
| **All Clusters** |  | -0.0051261 | 0.00044461 | -0.0060278 | -0.0042244 | 1.22E-13 |
| **Precentral** | Left | -0.0030003 | 0.0003537 | -0.0037176 | -0.0022829 | 4.15E-10 |
| **Precentral** | Right | -0.0046135 | 0.0004594 | -0.0055452 | -0.0036818 | 5.55E-12 |

Table A-22. Cluster-level associations between TARC and mean kurtosis within white matter. Mean diffusion values were extracted from significant clusters identified in voxel-wise analyses and entered into regression models controlling for age, sex, and education. β coefficients represent standardized effects. WM, white matter; MK, mean kurtosis.

| **Region** | **Hemisphere** | **Beta** | **SE** | **CI low** | **CI high** | **P** |
| --- | --- | --- | --- | --- | --- | --- |
| **All Clusters** |  | -0.0035073 | 0.00035988 | -0.0042372 | -0.0027774 | 1.23E-11 |
| **Anterior corona radiata** | Left | -0.0023851 | 0.00034918 | -0.0030933 | -0.0016769 | 5.48E-08 |
| **Anterior corona radiata** | Right | -0.0021795 | 0.00034032 | -0.0028697 | -0.0014893 | 2.01E-07 |
| **Body of corpus callosum** |  | -0.0029865 | 0.00042758 | -0.0038537 | -0.0021193 | 3.44E-08 |
| **Cingulate gyrus** | Left | -0.0057643 | 0.00075427 | -0.007294 | -0.0042346 | 4.81E-09 |
| **Cuneal cortex** |  | -0.005194 | 0.00061878 | -0.006449 | -0.0039391 | 5.36E-10 |
| **Genu of corpus callosum** |  | -0.0026771 | 0.0004586 | -0.0036072 | -0.001747 | 1.15E-06 |
| **Posterior corona radiata** | Right | -0.002947 | 0.00040992 | -0.0037783 | -0.0021156 | 1.86E-08 |
| **Posterior limb of internal capsule** | Left | -0.003104 | 0.0004119 | -0.0039393 | -0.0022686 | 6.59E-09 |
| **Posterior limb of internal capsule** | Right | -0.0036972 | 0.00046221 | -0.0046346 | -0.0027598 | 1.68E-09 |
| **Superior corona radiata** | Left | -0.0021807 | 0.00035633 | -0.0029034 | -0.0014581 | 4.81E-07 |
| **Superior fronto-occipital fasciculus** | Right | -0.005379 | 0.00073946 | -0.0068787 | -0.0038793 | 1.44E-08 |

Table A-23. Cluster-level associations between TARC and radial kurtosis within gray matter. Mean diffusion values were extracted from significant clusters identified in voxel-wise analyses and entered into regression models controlling for age, sex, and education. β coefficients represent standardized effects. GM, gray matter; RK, radial kurtosis.

| **Region** | **Hemisphere** | **Beta** | **SE** | **CI low** | **CI high** | **P** |
| --- | --- | --- | --- | --- | --- | --- |
| **All Clusters** |  | -0.006092 | 0.00043509 | -0.0069744 | -0.0052096 | 3.88E-16 |
| **Paracingulate** | Left | -0.0042817 | 0.00048336 | -0.005262 | -0.0033014 | 1.43E-10 |

Table A-24. Cluster-level associations between TARC and radial kurtosis within white matter. Mean diffusion values were extracted from significant clusters identified in voxel-wise analyses and entered into regression models controlling for age, sex, and education. β coefficients represent standardized effects. WM, white matter; RK, radial kurtosis.

| **Region** | **Hemisphere** | **Beta** | **SE** | **CI low** | **CI high** | **P** |
| --- | --- | --- | --- | --- | --- | --- |
| All Clusters |  | -0.0053286 | 0.0004729 | -0.0062877 | -0.0043695 | 2.33E-13 |
| Genu of corpus callosum |  | -0.0038805 | 0.00069193 | -0.0052838 | -0.0024772 | 2.32E-06 |
| Posterior corona radiata L | Left | -0.0049949 | 0.00064704 | -0.0063072 | -0.0036827 | 3.82E-09 |
| Superior corona radiata |  | -0.0088325 | 0.00105151 | -0.0109651 | -0.0067 | 5.27E-10 |

##### 1.5.2. Eotaxin-3

Table A-25. Cluster-Based associations between Eotaxin-3 and mean diffusivity within gray matter. Mean diffusion values were extracted from significant clusters identified in voxel-wise analyses and entered into regression models controlling for age, sex, and education. β coefficients represent standardized effects. GM, gray matter; CI, confidence interval.

| **Region** | **Hemisphere** | **Beta** | **SE** | **CI low** | **CI high** | **P** |
| --- | --- | --- | --- | --- | --- | --- |
| **All Clusters** |  | 0.00578947 | 0.00041778 | 0.00494217 | 0.00663678 | 5.31E-16 |
| **Precentral** | Right | 0.00722614 | 0.00146838 | 0.00424812 | 0.01020416 | 1.91E-05 |
| **Amygdala** | Left | 0.00776783 | 0.00190738 | 0.00389947 | 0.01163618 | 0.00024385 |
| **Cerebellum IV-V** | Right | 0.00397991 | 0.00096103 | 0.00203086 | 0.00592897 | 0.00019919 |
| **Frontal sup orb** | Left | 0.00575061 | 0.00090106 | 0.00392317 | 0.00757805 | 2.15E-07 |
| **Parietal inf** | Right | 0.0068495 | 0.00137675 | 0.00405733 | 0.00964167 | 1.62E-05 |

Table A-26. Cluster-Based associations between Eotaxin-3 and mean diffusivity within white matter. Mean diffusion values were extracted from significant clusters identified in voxel-wise analyses and entered into regression models controlling for age, sex, and education. β coefficients represent standardized effects. WM, white matter; CI, confidence interval.

| **Region** | **Hemisphere** | **Beta** | **SE** | **CI low** | **CI high** | **P** |
| --- | --- | --- | --- | --- | --- | --- |
| **All Clusters** |  | 0.00408094 | 0.0003315 | 0.00340862 | 0.00475325 | 1.83E-14 |
| **Anterior thalamic radiation** | Right | 0.00395152 | 0.00050351 | 0.00293036 | 0.00497267 | 2.62E-09 |
| **Inferior fronto-occipital fasciculus** | Right | 0.00367494 | 0.00064707 | 0.00236261 | 0.00498726 | 1.86E-06 |

Table A-27. Cluster-Based associations between Eotaxin-3 and radial diffusivity within gray matter. Mean diffusion values were extracted from significant clusters identified in voxel-wise analyses and entered into regression models controlling for age, sex, and education. β coefficients represent standardized effects. GM, gray matter; CI, confidence interval.

| **Region** | **Hemisphere** | **Beta** | **SE** | **CI low** | **CI high** | **P** |
| --- | --- | --- | --- | --- | --- | --- |
| **All Clusters** |  | 0.00610377 | 0.00044601 | 0.00519923 | 0.00700831 | 7.77E-16 |
| **Precentral** | Left | 0.00807237 | 0.0017855 | 0.00445121 | 0.01169354 | 6.43E-05 |
| **Amygdala** | Left | 0.00744758 | 0.00194841 | 0.00349602 | 0.01139914 | 0.00050453 |
| **Cerebellum IV-V** | Right | 0.0045066 | 0.0010173 | 0.00244341 | 0.00656979 | 8.45E-05 |
| **Frontal sup** | Left | 0.00614271 | 0.00104321 | 0.00402699 | 0.00825844 | 9.80E-07 |
| **Parietal inf** | Right | 0.00734997 | 0.00160629 | 0.00409227 | 0.01060767 | 5.45E-05 |

Table A-28. Cluster-Based associations between Eotaxin-3 and radial diffusivity within white matter. Mean diffusion values were extracted from significant clusters identified in voxel-wise analyses and entered into regression models controlling for age, sex, and education. β coefficients represent standardized effects. WM, white matter; CI, confidence interval.

| **Region** | **Hemisphere** | **Beta** | **SE** | **CI low** | **CI high** | **P** |
| --- | --- | --- | --- | --- | --- | --- |
| **All Clusters** |  | 0.00481186 | 0.00044688 | 0.00390554 | 0.00571818 | 8.30E-13 |
| **Frontal mid** | Right | 0.00469469 | 0.00064812 | 0.00338023 | 0.00600915 | 1.58E-08 |

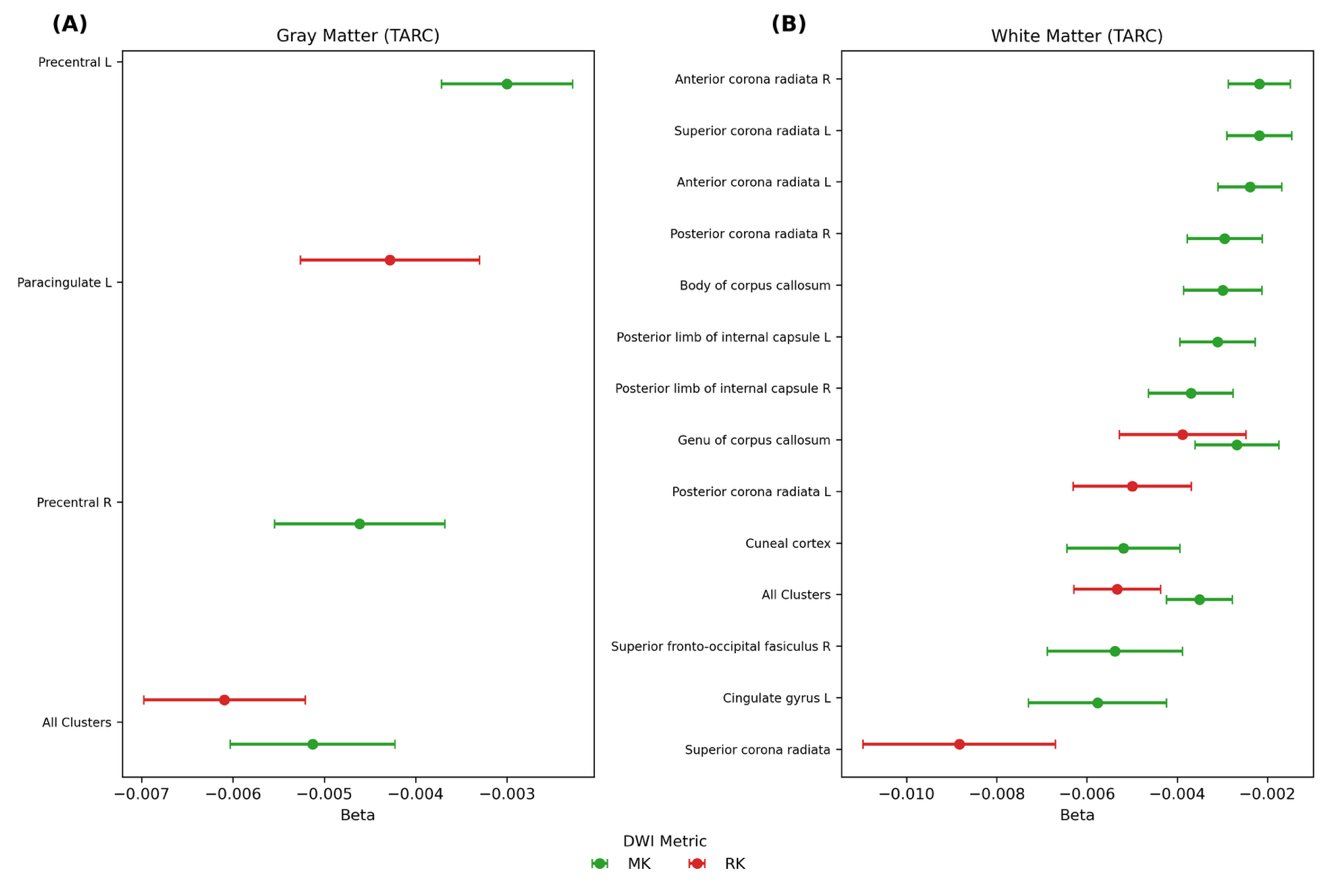

Figure A-3. **Cluster-based ROI associations between TARC and diffusion MRI metrics.** Forest plots showing cluster-level associations between plasma TARC and diffusion MRI metrics. Points represent standardized β estimates and horizontal bars indicate 95% confidence intervals from regression models adjusted for age, sex, and education. (A) Gray matter; (B) white matter. MK (green) and RK (red) are shown. The dashed line indicates β = 0. Clusters were defined from voxel-wise analyses surviving cluster-level FWE correction (p < 0.05).
